# T cell response to intact SARS-CoV-2 includes coronavirus cross-reactive and variant-specific components

**DOI:** 10.1101/2022.01.23.22269497

**Authors:** Lichen Jing, Xia Wu, Maxwell P. Krist, Tien-Ying Hsiang, Victoria L. Campbell, Christopher L. McClurkan, Sydney M. Favors, Lawrence A. Hemingway, Charmie Godornes, Denise Q. Tong, Stacy Selke, Angela C. LeClair, Chu-Woo Pyo, Daniel E. Geraghty, Kerry J. Laing, Anna Wald, Michael Gale, David M. Koelle

## Abstract

SARS-CoV-2 provokes a brisk T cell response. Peptide-based studies exclude antigen processing and presentation biology and may influence T cell detection studies. To focus on responses to whole virus and complex antigens, we used intact SARS-CoV-2 and full-length proteins with DC to activate CD8 and CD4 T cells from convalescent persons. T cell receptor (TCR) sequencing showed partial repertoire preservation after expansion. Resultant CD8 T cells recognize SARS-CoV-2-infected respiratory cells, and CD4 T cells detect inactivated whole viral antigen. Specificity scans with proteome-covering protein/peptide arrays show that CD8 T cells are oligospecific per subject and that CD4 T cell breadth is higher. Some CD4 T cell lines enriched using SARS-CoV-2 cross-recognize whole seasonal coronavirus (sCoV) antigens, with protein, peptide, and HLA restriction validation. Conversely, recognition of some epitopes is eliminated for SARS-CoV-2 variants, including spike (S) epitopes in the alpha, beta, gamma, and delta variant lineages.

## Introduction

The acquired immune response to SARS-CoV-2 can limit infection, as shown by protection from re-infection (1) and the efficacy of vaccines (2). While antibodies can prevent infection and disease, T cell depletion studies of convalescent or vaccinated animals strongly suggest active roles for T cells (3). Many studies have defined regions of the predicted SARS-CoV-2 proteome that can activate T cells in blood (4, 5). This work typically uses single or pooled peptides, or peptide-based reagents such as HLA multimers. Some workflows result in cell death that limit follow-up, while others, such as activation-induced marker (AIM)-based cell sorting (6) allow recovery of live cells for downstream work. Taken together, peptide-based studies provide a large thesaurus of reactive peptides, and in some cases, relevant HLA restricting alleles and TCR sequences. However, validation of reactivity with whole virus or complex antigens is less frequently reported.

The present report used PBMC from a cohort of coronavirus-19 (COVID-19) convalescent persons (7–10) to begin to study T-cell responses to SARS-CoV-2 in the contexts of direct- and cross-presentation of complex viral antigens. Validation with proteome-covering full-length protein and peptide sets allowed definition and confirmation of many individual epitopes.

Expanded responder cell populations also permit detailed study of HLA restriction, functional avidity, cross-recognition of sCoV, and recognition of circulating SARS-CoV-2 variants including variants being monitored (VBM) and variants of concern (VOC).

Estimates of the overall magnitude of the CD4 and CD8 T cell response to SARS-CoV-2, as a percent of circulating PBMC at times soon after clinical recovery are around the 0.5-1% level based on the summation of peptide reactivities (4, 11, 12). We were interested in benchmarking peptide-based estimates for SARS-CoV-2 to the levels of reactivity to whole viral antigen, and in comparing epitope specificities determined using peptide sets and more complex antigens. It is well established that T cell clonotypes show cross-reactivity to unrelated peptide epitopes presented by a single HLA allele, for example cross-recognition of disparate influenza and EBV peptides with HLA A*02:01 (13), pathogenic cross-recognition of influenza and self-epitopes (14), and potentially beneficial cross-reactivity between viruses and tumor antigens (15). The same TCR can also recognize peptides presented by divergent HLA alleles, for example the alloreactivity of herpes simplex virus (HSV)- or EBV-specific T cells against HLA-mismatched antigen presenting cells (APC) bearing endogenous peptides (16, 17). Reactivities detected solely using SARS-CoV-2 peptide sets could include such cross-reactivity. The use of complex SARS-CoV-2 antigens to enrich responses also incorporates antigen processing and presentation steps, such as proteosomal cleavage, peptide transport into the endoplasmic reticulum and trimming during HLA loading, that can be influenced by flanking sequences within proteins, as well as potentially incorporating post-translational modifications and cryptic ORFs absent from peptide sets (18, 19). We also studied recognition of whole SARS-CoV-2 at the effector stage: CD8 T cell assays used infected bronchial epithelial cells HLA-matched to CD8 effector T cells, while CD4 T cell readouts used SARS-CoV-2 antigen and appropriate APC. Importantly, we also checked if T cells enriched using simpler peptide or protein antigens could recognize whole virus.

SARS-CoV-2 shares genomic structure and sequence motifs with sCoV. Cross-reactivity with sCoV has been described (20) but is less studied in the whole virus context. SARS-CoV-2 also shows modest sequence variation, and variants and deletions emerge within-subject in immune suppressed persons (21), and within populations (22). In the present report, we use culture-amplified T cell responders to probe recognition of sCoV and variant SARS-CoV-2 T cell epitopes.

## Results

### Subjects and specimens

We studied 26 specimens from 12 subjects (Supplementary Table 1) with COVID-19 in March or April 2020, prior to detection of VOCs in the study region (23). Each subject reported SARS-CoV-2 RNA detection, positive serum or plasma anti-S domain 1 and anti-N IgG, and SARS-CoV-2 neutralizing antibody (nAb) titer ≥ 1:40 (7). Median age was 56 (range 32 to 72) and sex was balanced. Amongst 4 hospitalized subjects, 3 required intensive care. Median illness duration was 16 days (range 4 to 32). PBMC were obtained a median of 138 days after recovery (range 31 to 256). We used several methods to enrich SARS-CoV-2-specific T cells for detailed studies (outlined in Supplementary Fig. 1).

### PBMC T cell responses to whole SARS-CoV-2 antigen

SARS-CoV-2 WA1 antigen was prepared from infected cells, rather than from purified virions, to include non-structural proteins (NSPs) (24). We used dual expression of CD137 and CD69 for analytic AIM and to sort cells for expansion (Fig. 1) (25). We observed a median of 0.16% (range 0.089% to 0.33%) of CD4 T cells were AIM (+) (n=5, Supplementary Table 2). Responses to mock antigen were low (median 0.026%, range 0.009%-0.061%). Responses to viral antigen were higher than to mock antigen (p =0.0078 by pair-wise analysis); the median net response was 0.13%. Absolute and net responses to whole SARS-CoV-2 antigen in HD PBMC were low, with no significant net virus-specific signal (Supplementary Fig. 2B). We sorted a median of 389 AIM (+) CD4 T cells (machine counts, range 375 - 2,138) per specimen from the 5 COVID-19 convalescent subjects (Supplementary Table 2).

**Fig. 1.**
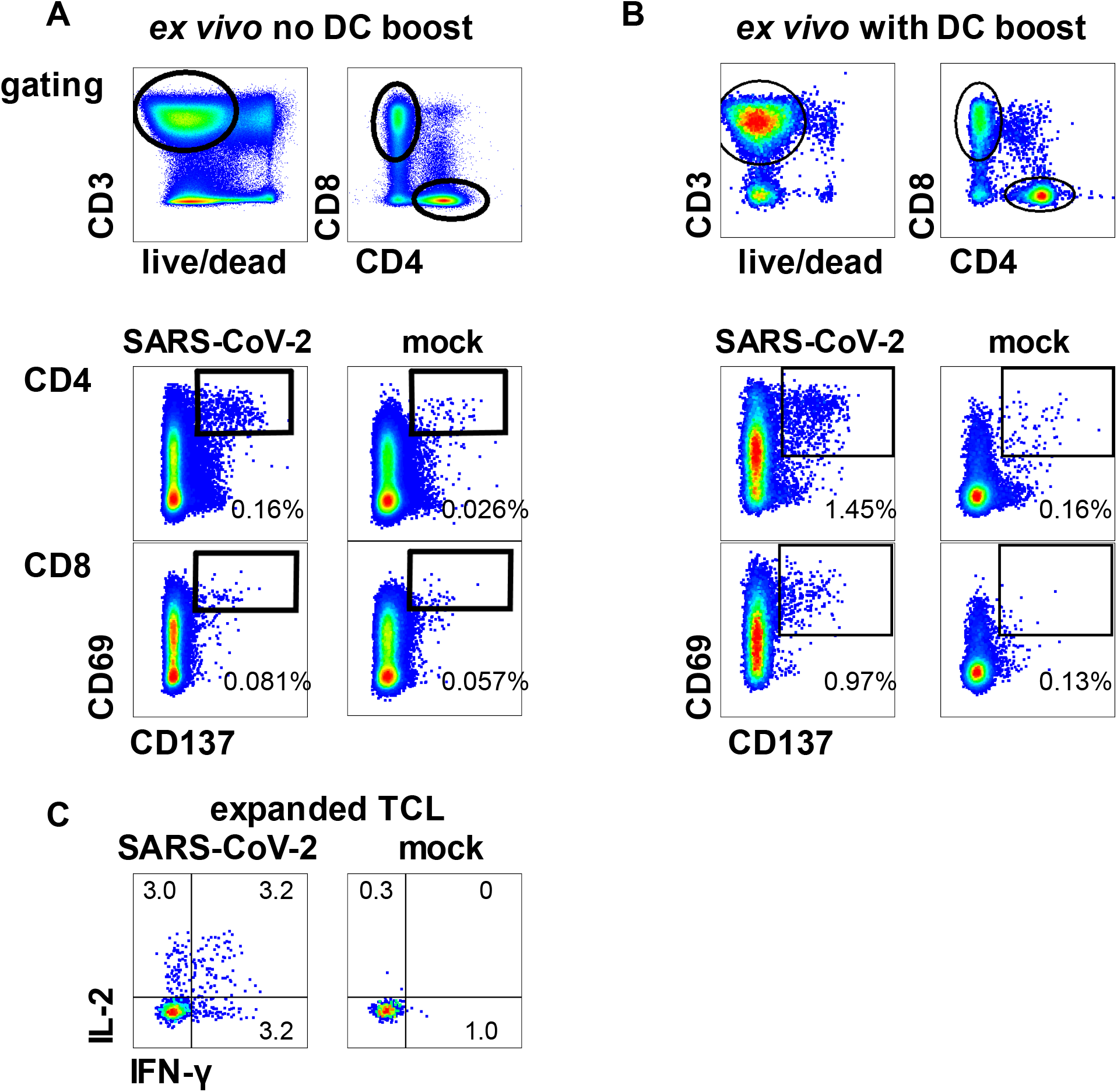
AIM detection and enrichment of SARS-CoV-2 specific T cells in response to whole virus. A: PBMC from participant W003 incubated with inactivated cell-associated SARS-CoV-2 or mock antigen. Gating scheme at top. Lower panels show expression of activation markers CD137 and CD69 in response to 18 hour stimulation amongst CD4 or CD8 T cells. B: Similar layout for subject W005, specimen 1 PBMC stimulated with autologous moDC pre-treated with SARS-CoV-2 or mock antigen. Gating scheme at top. For both stimulation methods, numbers are percentages of gated T cells expressing dual activation markers. C: CD69/CD137 positive CD4 T cells from the pathway in part A: were expanded and tested for reactivity with inactivated cell-associated SARS-CoV-2 or mock antigen. Gated, live, responder, CD3+/CD4+/CD8-cells are shown. Numbers are percent of cells accumulating the indicated cytokines.

After expansion with a generalized mitogenesis protocol, T cell line (TCL) functional enrichment was measured as reactivity to whole killed SARS-CoV-2. Autologous PBMC used as APC were dump-gated and IFN-γ and IL-2 expression used to enumerate SARS-CoV-2-specific CD4 T cells (8, 11). We observed robust TCL enrichment of virus-reactive CD4 T cells. For example, participant W003 TCL had net 8.14% of cells responding to whole SARS-CoV-2 (Fig. 1C), a 61- fold enrichment over the PBMC CD4 T cell AIM signal. Enrichment ranged from 41- to 201-fold for the 5 subjects (Supplementary Table 2).

Both monocytes in fresh PBMC and moDC can process and present antigens to CD4 T cells. Higher levels of PBMC CD4 T cell activation were noted when autologous DC were used as initial APC (Fig. 1B) compared to direct addition of killed virus to PBMC. Background signal for mock antigen also slightly increased (median 0.16%, range 0.11%-0.47%). The median net proportion of AIM (+) CD4 T cells was 0.66% (range 0.35% to 2.43%) across 12 samples. Use of moDC allowed the sorting of a median of 11,320 activated CD4 T cells per specimen (range 8,190 to 19,860), from which 20% were culture-expanded (Supplementary Table 2) and 80% used for *ex vivo* TCR sequencing.

In addition to presentation to CD4 T cells, moDC can cross-present inactivated antigen to CD8 T cells. Using moDC, we observed specific CD8 T cell activation as measured by CD69 and CD137 co-expression (Fig. 1B). Negative control antigen was immunologically silent. We did not detect PBMC CD8 T cell responses in HD (Supplementary Fig. 2). As with CD4 T cells, higher net proportions of AIM (+) CD8 T cells were noted with moDC (n = 12, median 0.95%, range 0.27 to 1.62%) than without (n = 5, median 0.033%, range 0.013%-0.29%) (Supplementary Fig. 2B, Supplementary Table 2). A median of 842 CD8 T cells (range, 411 - 2,611) were expanded per specimen, while 80% of the AIM-sorted cell populations were TCR-sequenced.

Two subjects were studied using moDC at five time points each between 32 to 256 days after recovery from COVID-19. Decreasing CD4 and CD8 AIM (+) abundance was noted over time (Supplementary Table 2). Subject W005 had consistently higher CD4 than CD8 T cell responses, with the ratio of activated cells ranging from 1.54 to 3.15. A reciprocal pattern was seen in subject W012, with consistent CD4/CD8 ratios of AIM (+) cells < 1.

### TCR repertoire tracking during T cell expansion

We used the moDC workflow to present whole SARS-CoV-2 antigen (above), and analyzed AIM (+) CD4 and CD8 T cells from three subjects at two time points each. A portion of AIM (+) cells were directly sequenced, and a portion sequenced after expansion, with a genomic DNA-based method (26). A median of 6.75% of CDR3 aa sequences detected *ex vivo* were found in expanded cultures. Reciprocally, a median of 16.75% of productive CDR3 aa sequences in the expanded cultures were detected in the corresponding ex vivo samples (representative data, Supplementary Fig. 3A). CDR3 abundances showed excellent agreement for AIM (+) cells after one vs two expansions (representative data, Supplementary Fig. 3B). A possible factor contributing to repertoire differences *ex vivo* vs. post-expansion is poor recovery of DNA from small *ex vivo* specimens. Amongst the 12 *ex vivo* samples, the median number of productive CDR3 gene rearrangements reported was 11.6% (range, 4.9-21.7%) of the sorting cell counts.

### Alternative generation of polyclonal SARS-CoV-2-reactive T cells

As an alternative approach to retains an antigen processing requirement, SARS-CoV-2 proteins were expressed in COS-7 or HeLa cells. These were harvested, inactivated, and added to PBMC without (COS-7) or with (HeLa) autologous moDC. Small increments in AIM-positive CD4 or CD8 T cells were detectable compared to mock antigen (representative data, Supplementary Fig. 4A). Finally, CD8 or CD4 T cells proliferating (representative data, Supplementary Fig. 5A) to SARS-CoV-2 peptide pools were sorted and expanded.

### CD8 T cell recognition of infected cells

CD8 T cell recognition typically requires HLA matching. HBEC3-KT-A are permissive for SARS-CoV-2 replication and are HLA A*03:01 (+) (Supplementary Table 3). For effector CD8 T cells we stimulated PBMC from HLA A*03:01 (+) participant W003 with autologous DC loaded with S protein-expressing HeLa cells, sorted AIM (+) CD8 T cells, expanded them, and showed they recognized S protein in the HLA A*03:01 context (Supplementary Fig. 6A). A reactive peptide, S 377-389, was found using peptide matrices (Supplementary Fig. 6B) containing the HLA A*03:01-restricted epitope S 378-386 (27). A fluorescent A*03:01-S 378-386 tetramer allowed further enrichment of CD8 T cells (Supplementary Fig. 6C). We then examined if S protein could be processed and presented by virus-infected cells. CD8 T cells specifically recognized SARS-CoV-2-infected HBEC3-KT-A cells (Fig. 2A). Specificity control HLA A*03:01-restricted, tetramer-enriched CD8 T cells, specific for an unrelated polyomavirus epitope in MCPyV (28), were active when tested with polyomavirus peptide-pulsed HBEC3-KT-A positive controls (Fig. 2B) but did not recognize SARS-CoV-2-infected target cells (Fig. 2A).

**Fig. 2.**
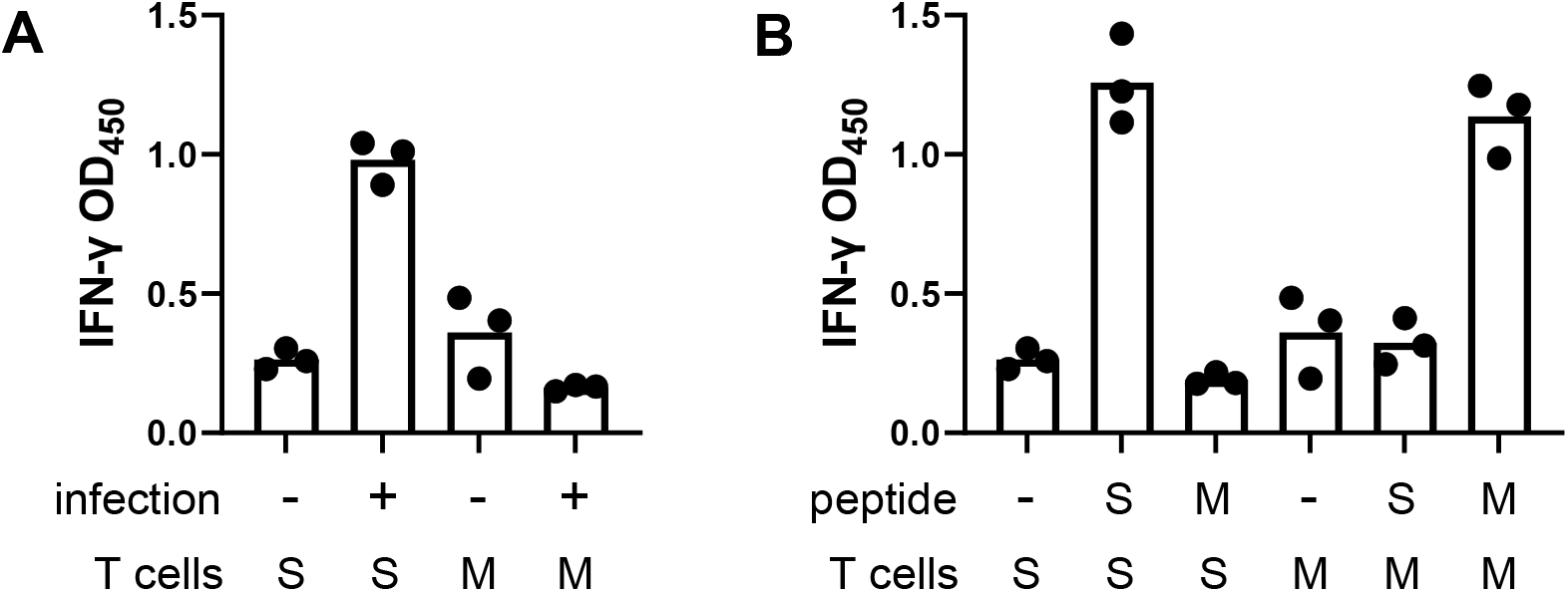
Recognition of infected respiratory epithelial cells by SARS-CoV-2-specific CD8 T cells. A: HBEC3-KT-A cells with or without infection with SARS-CoV-2 were co- cultivated with tetramer-enriched, HLA A*03:01-restricted, S 378-386-specific CD8 T cells and activation measured by IFN-γ secretion. S: S-specific T cells. M: control MCPyV-specific CD8 T cells. B: Both T cell populations specifically recognized HBEC3- KT-A cells treated with relevant viral peptide.

### CD8 T cell responses to full-length SARS-CoV-2 proteins

CD8 T cell target diversity was studied using whole virus stimulation followed by SARS-CoV-2 proteome-wide scans using subject-specific HLA A and B aAPC panels. A complex response was observed for subject W005, hospitalized in the ICU and studied 32 days after recovery, by incorporating DC boosting. This subject had an HLA B*15:02-restricted response to NSP2, HLA A*11:01- and B*15:02- restricted responses to nucleoprotein, and an HLA B*51:01-restricted response to ORF3A (Fig. 3A). Multiple proteins were positive for subject W012, while subject W001, studied using moDC, only showed responses to ORF9B (summarized in Fig. 4A). Even when moDC were not used, the aAPC-proteome panels were successful for bulk AIM-enriched CD8 TCL enriched with whole virus. For example, subject W010 had a single HLA A*01:01-restricted response detected for NSP3 (Fig. 3B). This was confirmed at the peptide level (Fig. 3C), allowing tetramer enrichment (Fig. 3D) and use of tetramer-enriched cells confirmed as ORF- and peptide-reactive (Fig. 3E) to determine functional avidity (Fig. 3F).

**Fig. 3.**
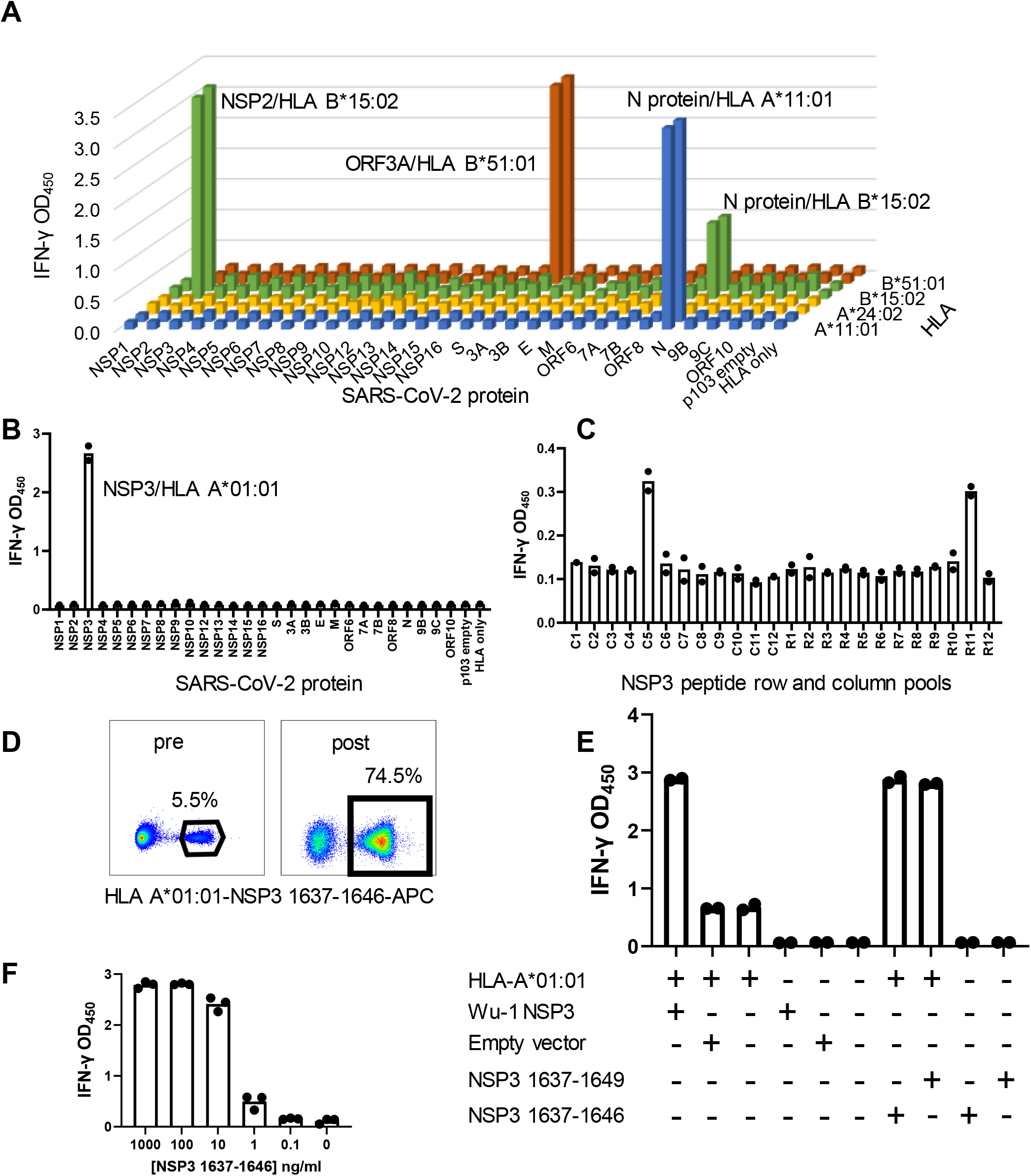
SARS-CoV-2 CD8 T cell antigens and epitopes from PBMC stimulation with whole SARS-CoV-2 antigen. A: Subject W005, specimen 1 CD8 TCL were assayed with aAPC expressing each SARS-CoV-2 protein and relevant HLA A and B. Four reactivities were noted. B: Subject W010 CD8 TCL is reactive with HLA A*01:01 aAPC co-transfected with NSP3. For A: and B: negative controls at right. C: CD8 TCL from B: assayed against column (C) and row (R) pooled NSP3 peptides with autologous EBV-LCL as APC. D: Tetramer stain of CD8 TCL before and after sorting and expansion of tetramer-positive cells. Percentages of tetramer-positive cells shown E: Reactivity of tetramer-enriched cells for aAPC transfected with the indicated plasmids or treated with the indicated peptides. F: Dose-response for HLA A*01:01 aAPC with the indicated concentrations of NSP3 1637-1646. Duplicate or triplicate IFN-γ release assays show raw data as bars (A) or dots for each value and means as bars (B, C, E, F).

**Fig. 4.**
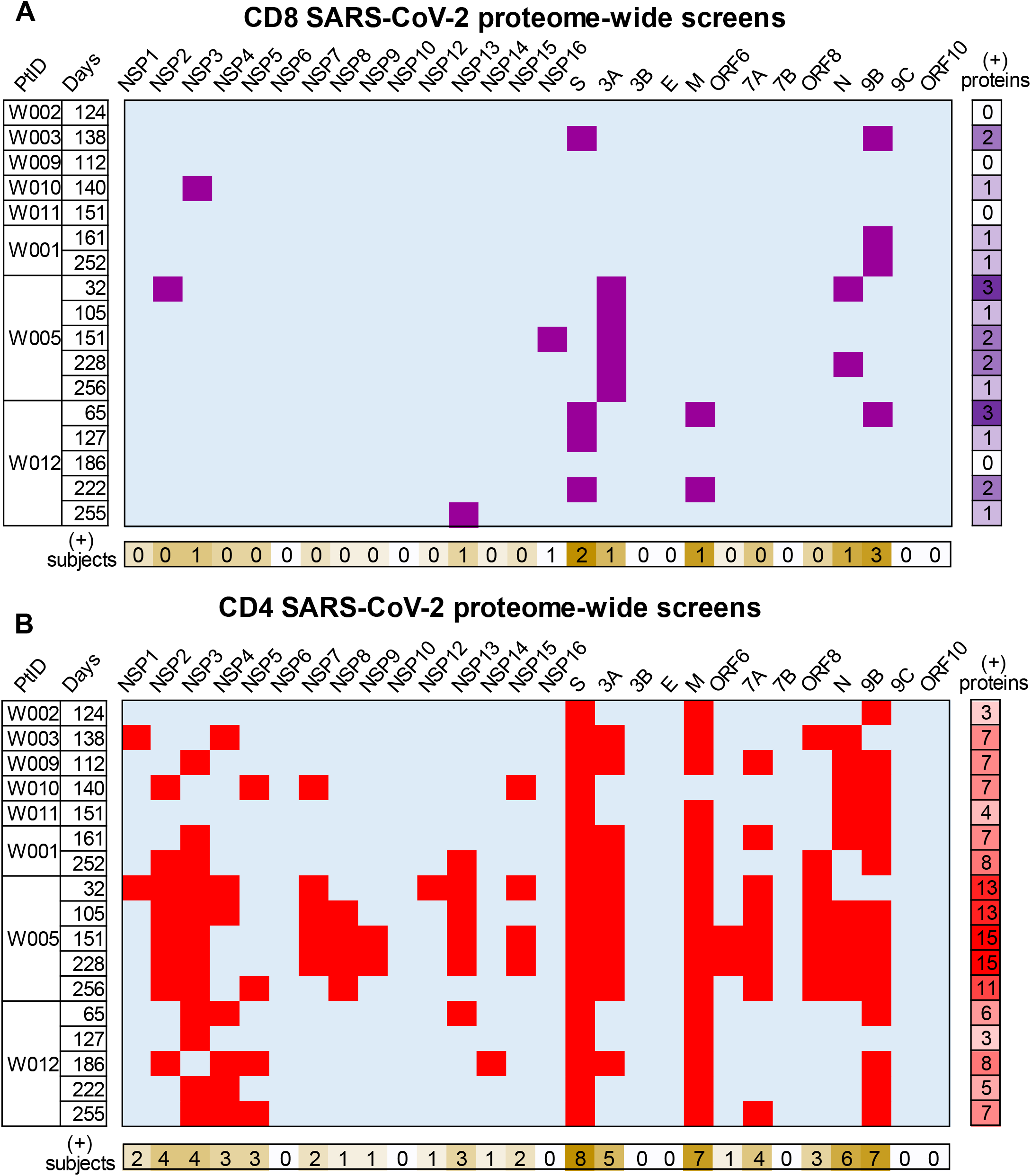
Summary of SARS-CoV-2 proteome-level T cell reactivity for PBMC after COVID-19 illness, using AIM enrichment with whole viral antigen. Rows indicate donors and days between recovery from illness and PBMC sampling. Upper 5 rows were studied without DC boosting; lower 12 rows used this procedure. Columns are individual SARS-CoV-2 proteins. A: CD8 TCL scoring positive (purple). B: CD4 TCL scoring positive (red). At right are number of proteins recognized and at bottom are number of subjects with reactivity at one or more time points.

### CD4 T cell responses to full-length SARS-CoV-2 proteins

AIM-sorted, expanded CD4 TCL, enriched with or without DC boosting, were similarly tested proteome-wide. Each SARS-CoV-2 ORF or cleaved polypeptide product of ORFs polypeptide (PP)1a and PP1ab(29) was expressed (30). Readout assays included Th1 cytokine ICS (supplementary Fig. 7A), proliferation (supplementary Fig. 7B), and IFN-γ secretion (supplementary Fig. 7C). Across 17 specimens from 8 persons, a median of 7 proteins were recognized per specimen (range, 3 to 15) (summarized in Fig. 4B). The SARS-CoV-2 proteins recognized in the highest percentage of participants were S, membrane (M), nucleoprotein (N), and ORF9B (Fig. 4B). Reactivity to ORF3A, ORF7A, NSP2, and NSP3 were also noted in 50% or more of participants. Because DC were not used for all specimens and few participants were studied, conclusions about antigen breadth and dominance remain preliminary. We noted that over time, most within-participant protein-level responses were consistent. For example, participant W005 recognized NSP2, NSP13, and NSP16 at almost all time points (Fig. 4B).

### CD8 T cell epitopes

TCL initiated with diverse workflows (Supplementary Fig. 1) were used. moDC-whole virus stimulation of PBMC yielded peptide epitopes (examples, Supplementary Fig. 8) to confirm ORF-level hits (Fig. 3A, Fig. 4A). PBMC stimulation with SARS-CoV-2 proteins with DC, followed by AIM sorting also yielded cultures that reacted with the relevant ORF and discrete peptides (example, Supplementary Fig. 4A, 4B), as did PBMC stimulation with pooled peptides followed by sorting of proliferated cells (Supplementary Fig. 5A, Supplementary Fig. 4C). Overall, integrated across CD8 TCL and eliminating redundancies, CD8 T cells reactive with 25 SARS-CoV-2 epitopes in the context of 8 HLA class I alleles were obtained (Supplementary Table 4).

### CD4 T cell epitopes

TCL initiated with several workflows were tested to define peptide epitopes. Importantly, when TCL were started with peptide pools (as in Supplementary Fig. 5A), the resultant TCL recognized whole virus lysate and full-length protein (Supplementary Fig. 5B) as well as peptides (Supplementary Fig. 5C), suggesting epitopes discovered with peptides are relevant to viral infection. IFN-γ and proliferation readouts corresponded (representative data, Supplementary Fig. 9) and we summarize epitopes defined with either ror both readouts. Cultures initiated with complex antigen, such as from participant W001 using moDC-assisted stimulation with whole virus, yielded multiple reactive peptides (representative data, Supplementary Fig. 10). Overall, CD4 T cell epitopes were detected in 13 SARS-CoV-2 proteins. Responses were diverse within-person. For specimen 1 from participant W001 (Supplementary Table 1), 55 epitopes were confirmed in 7 proteins, including 27 epitopes in S, 7 each in N and NSP3, 6 in ORF3A, 5 in M, 2 in ORF9B, and 1 in ORF7A (Supplementary Table 5). Similarly, in participant W005 studied one month after recovery, 65 CD4 T cell epitopes were identified in 10 proteins. Altogether, we observed 240 CD4 T cell peptide reactivities from 8 persons (Supplementary Table 5). These correspond to 172 unique SARS-CoV-2 peptides (Supplementary Table 5). Amongst these, we found 70 epitopes in S protein, 26 in N protein, and 23 in M protein, with fewer in ORFs 3A, 7A, 8, and 9, and NSPs 1, 2, 3, 4, 6, and 7. Peptides with CD4 T cell recognition by at least half the population studied included M 69-81, M 177-189, S aa 133-145, S 165-177, N 289-301, and N 349-361.

### CD4 T cell HLA restriction and functional avidity

Studies of CD4 T cell HLA restriction and estimated functional avidity (representative data, Supplementary Fig. 11) yielded HLA locus-level data on 118 of 123 (96%) peptides tested. Amongst these, 84 (72%) were HLA DR-restricted, 23 (20%) were HLA DQ-restricted, and 11 (8%) were HLA DP-restricted (Supplementary Table 5). HLA restriction at the locus level was generally identical if two participants recognized the same peptide, but there were exceptions. S protein aa 165-177 was HLA DP-restricted in 4 participants and HLA DR-restricted in 2. Allele-level restriction was determined using aAPC (representative data, Supplementary Fig. 11A, 11B). We observed presentation by 10 distinct HLA DRB1, DRB3, and DRB4 alleles (Supplementary Table 5). Some peptides in SARS-CoV-2 S, M, and N proteins were presented by both DRB1 and either DRB3 or DRB4 (representative data, Supplementary Fig. 11C) or dual DRB3 alleles (not shown). Functional avidity data were available for 124 CD4 T cell epitopes (representative data, Supplementary Fig. 11, summarized in Supplemental Table 5). Responses at 1 and 10 ng/ml peptide were noted for 4 and 10 epitopes, respectively. S 165-177 was particularly potent, with responses at 1 or 10 ng/ml for 5 of 6 subjects.

### Recognition of seasonal coronaviruses

Cross-recognition of SARS-CoV-2 and sCoV peptide has been documented (6, 20), but less is known about recognition of complex viral antigens. We found that polyclonal CD4 TCL each of three subjects studied, enriched from PBMC using whole SARS-CoV-2 antigen and moDC, cross-recognized either whole OC43 or whole 229E cell-associated virus (Fig. 5A). Control mock-infected virus preparations were negative. An additional CD4 TCL enriched using whole SARS-CoV-2 antigen without moDC also cross-recognized OC43. For subject W001, S proteins from both viruses were both antigenic (Fig. 5B), as were both homologs of a peptide in S. The HLA restricting allele was established as HLA DRB1*15:01 (Fig. 5C). Cross-recognition of an HLA DP-restricted peptide that is nearly identical in SARS-CoV-2, OC43, and HKU1 N proteins was also observed (Supplementary Fig. 12).

**Fig. 5.**
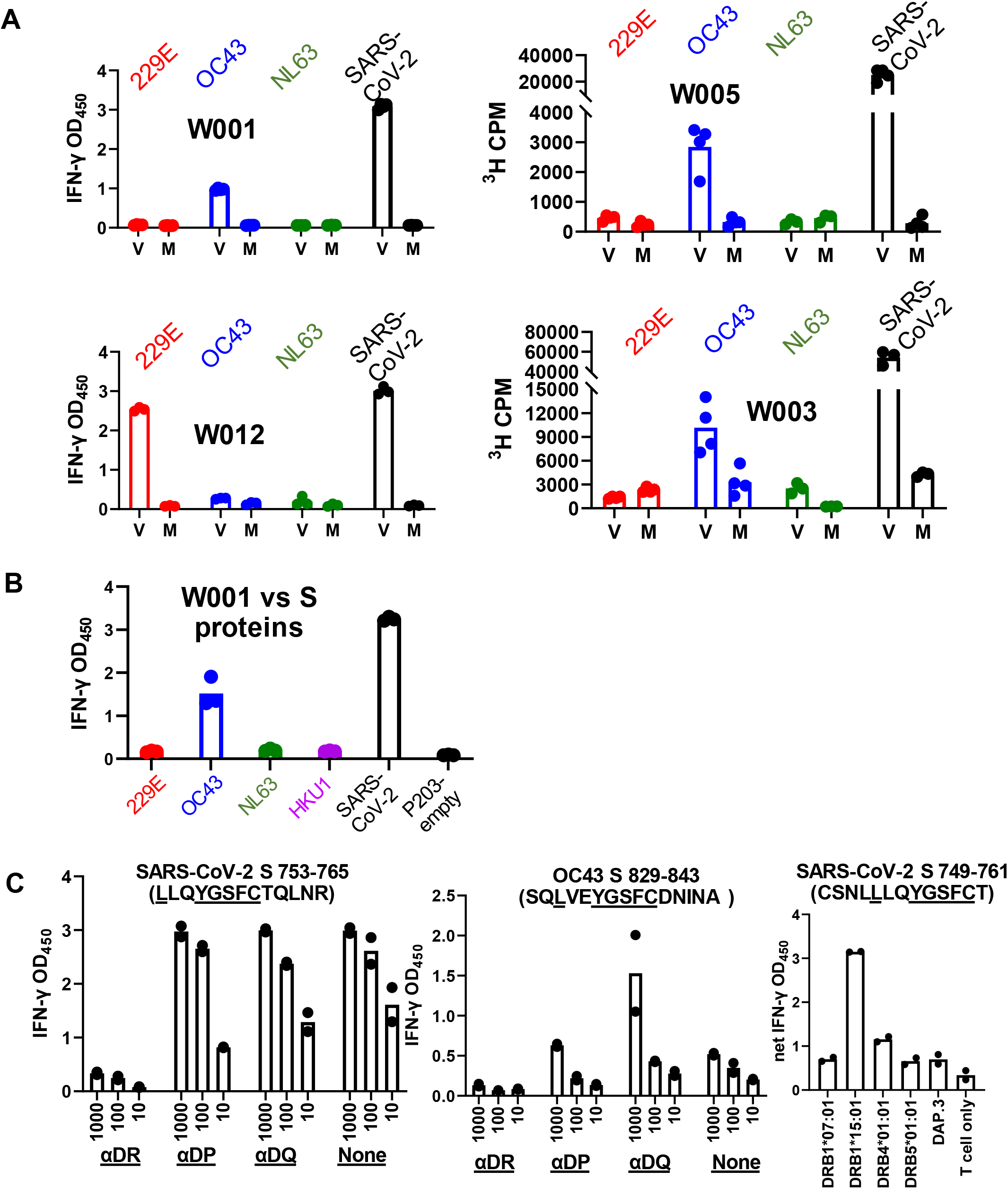
A: CD4 TCL from PBMC stimulated with moDC and whole SARS-CoV-2 (subjects W001, W005, W012) or PBMC stimulated with whole SARS-CoV-2 (subject W003) recognize whole SARS-CoV-2, and sCoVOC43 or 229E antigens (V), but not mock (M) antigens. B: CD4 TCL from subject W001 recognizes full-length S protein from OC43 but not empty vector control. C: CD4 TCL recognize homologous S peptides from SARS-CoV-2 and OC43 in an HLA DR- restricted fashion as indicated by inhibition with locus-specific mAb. An overlapping SARS-CoV- 2 peptide shows DRB1*15:01 restriction at right. Conserved AA are underlined.

### Recognition of SARS-CoV-2 variants

To choose variants for study we correlated SARS-CoV-2 T cell epitopes detected using Wu-1/WA1 reagents with variants both in early 2020 data and in early 2021 VBM, concentrating on the B.1.1.7, B.1.351, and P1 lineages. Polyclonal CD8 TCL were recovered from subject W004 using an S peptide pool (Supplementary Fig. 5A). Potent recognition of strain Wu-1 peptides 269-277 YLQ and 417-425 KIA, HLA A*02:01- restricted epitopes (27), was observed (Fig. 6). There was no recognition of variants with substitutions K417N and K417T which are present in the B.1.351 and P.1 lineages, respectively. Epitope-specific T cells enriched with tetramers (Supplementary Fig. 13A) detected full-length S processed by HLA A*02:01-transfected aAPC (Supplementary Fig. 13B), but failed to recognize S K417T or the full-length SARS-CoV-2 S variant from lineage B.1.351 bearing K417T. Responses to S from B.1.1.7, which does not have an aa417 substitution, were intact. Control CD8 T cells specific for S aa 269-277 YLQ, unchanged in these variants, were not affected (Supplementary Fig. 13B). We also observed loss of CD8 TCL reactivity to SARS-CoV-2 S F490S in the HLA A*29:02-restricted aa 489-497 epitope, to S K378N in the HLA A*03:01- restricted aa 378-386 epitope, and to M (membrane) T175M in the HLA A*11:01 aa 169-181 epitope (Supplementary Fig. 13, summarized in Table 1).

**Fig. 6.**
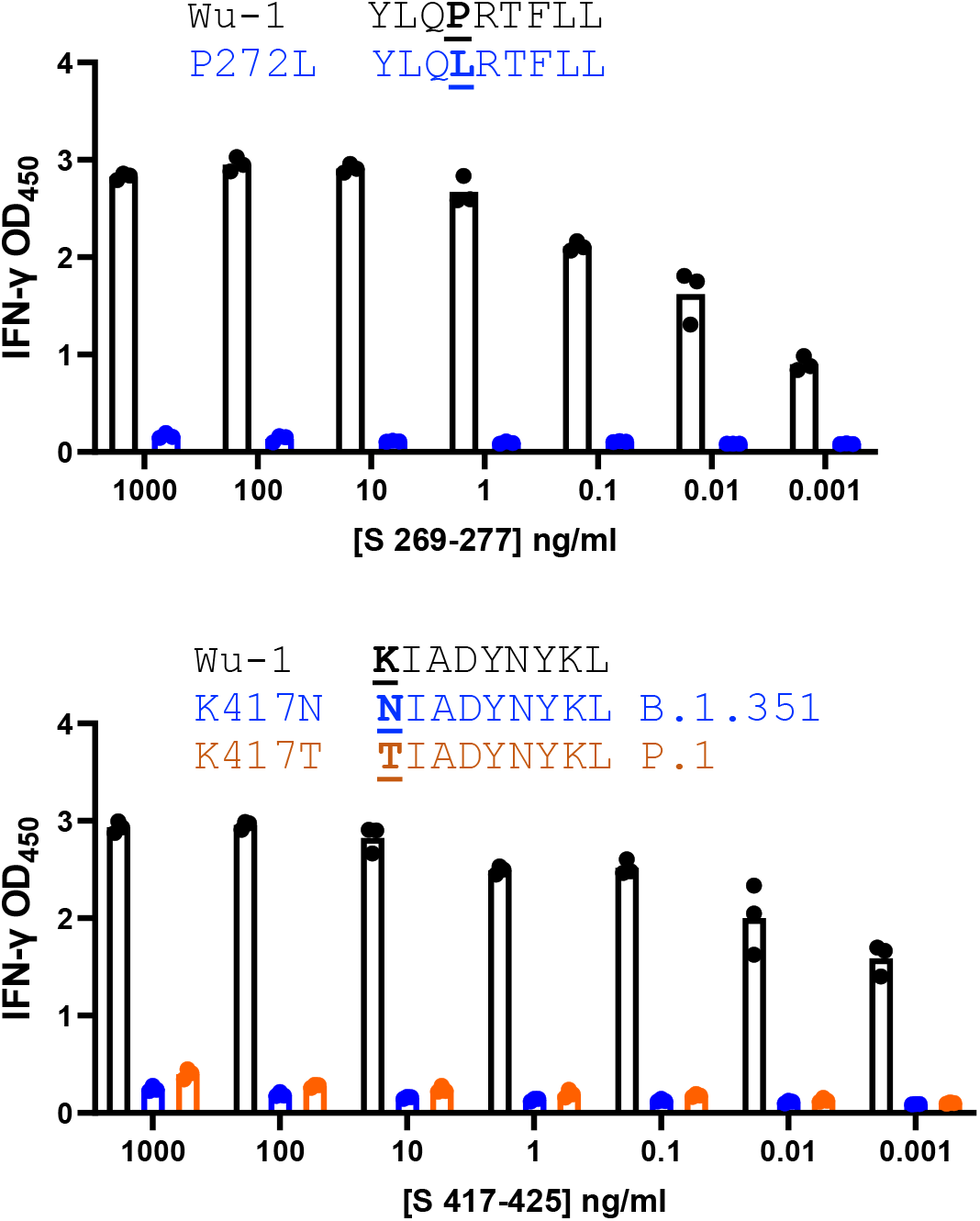
CD8 TCL recognition by SARS-CoV-2 variant peptides. aAPC transfected with HLA A*02:01 and treated with wild-type but not variant peptides were recognized by polyclonal CD8 TCL lines. Lineage B.1.351 is also known as beta and P.1 is also known as gamma).

**Fig. 7.**
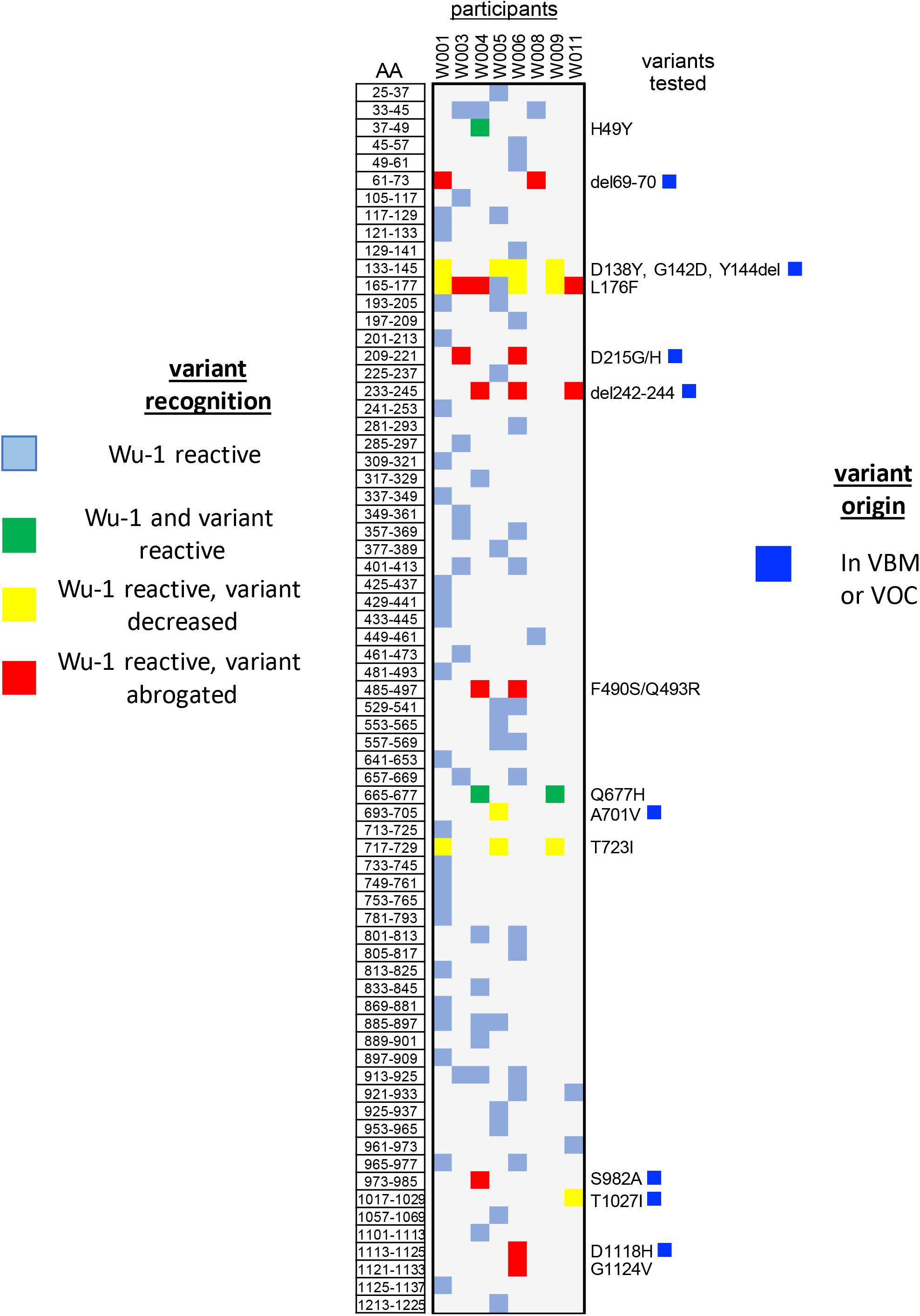
CD4 TCL recognition by SARS-CoV-2 variant peptides. Summary of recognition of S strain Wu-1 peptides and variants. Donors indicated at top. Each Wu-1 peptide recognized by one or more subject is numbered at left. Coordinates and AA substitutions or deletions of variants tested are listed at right. Variants in WHO VOC indicated with blue square. Color codes at left summarize level of recognition of variant peptide by each TCL. VBM = variants being monitored, VOC = variants of concern per US CDC October 2021.

**Table 1.**
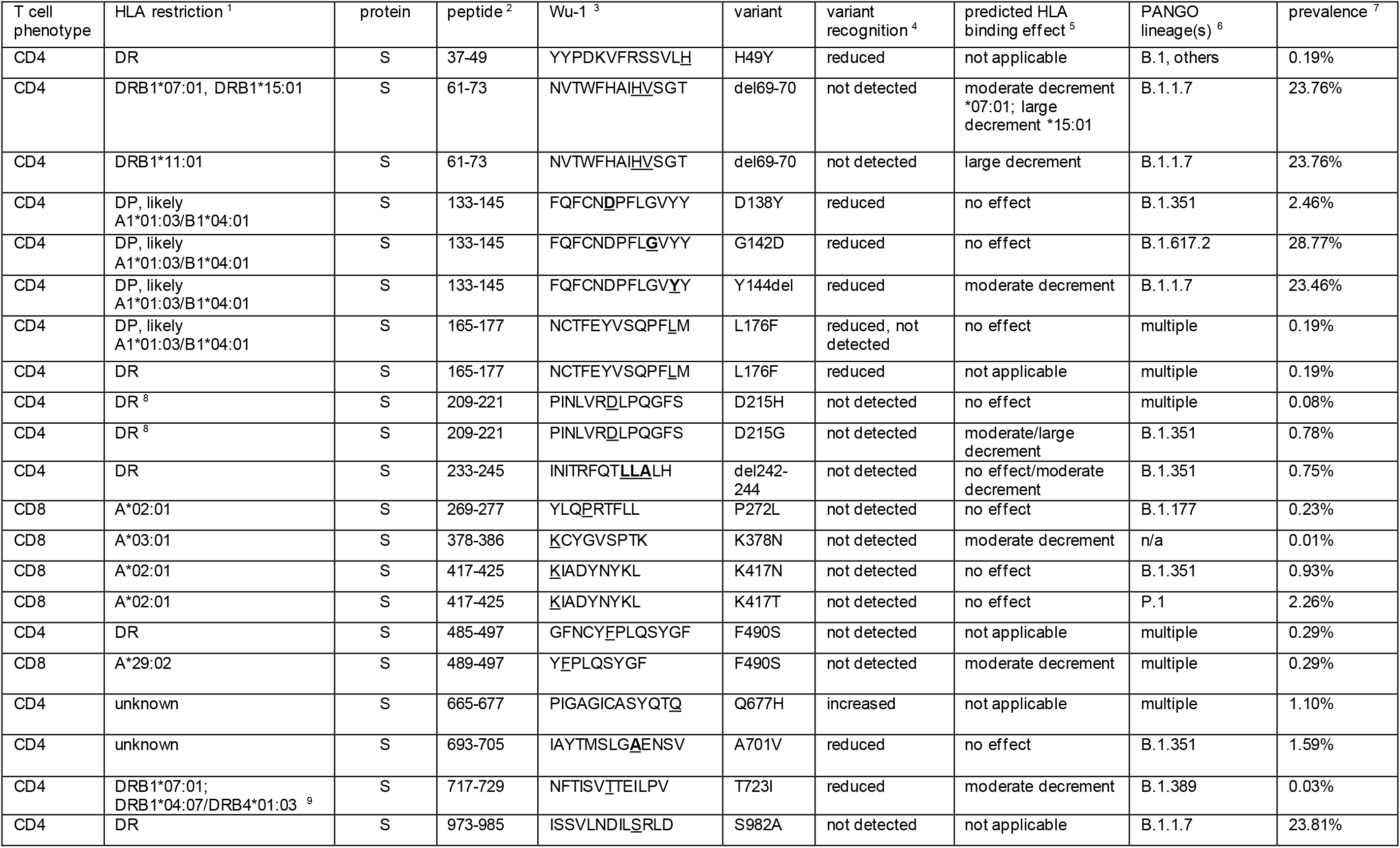
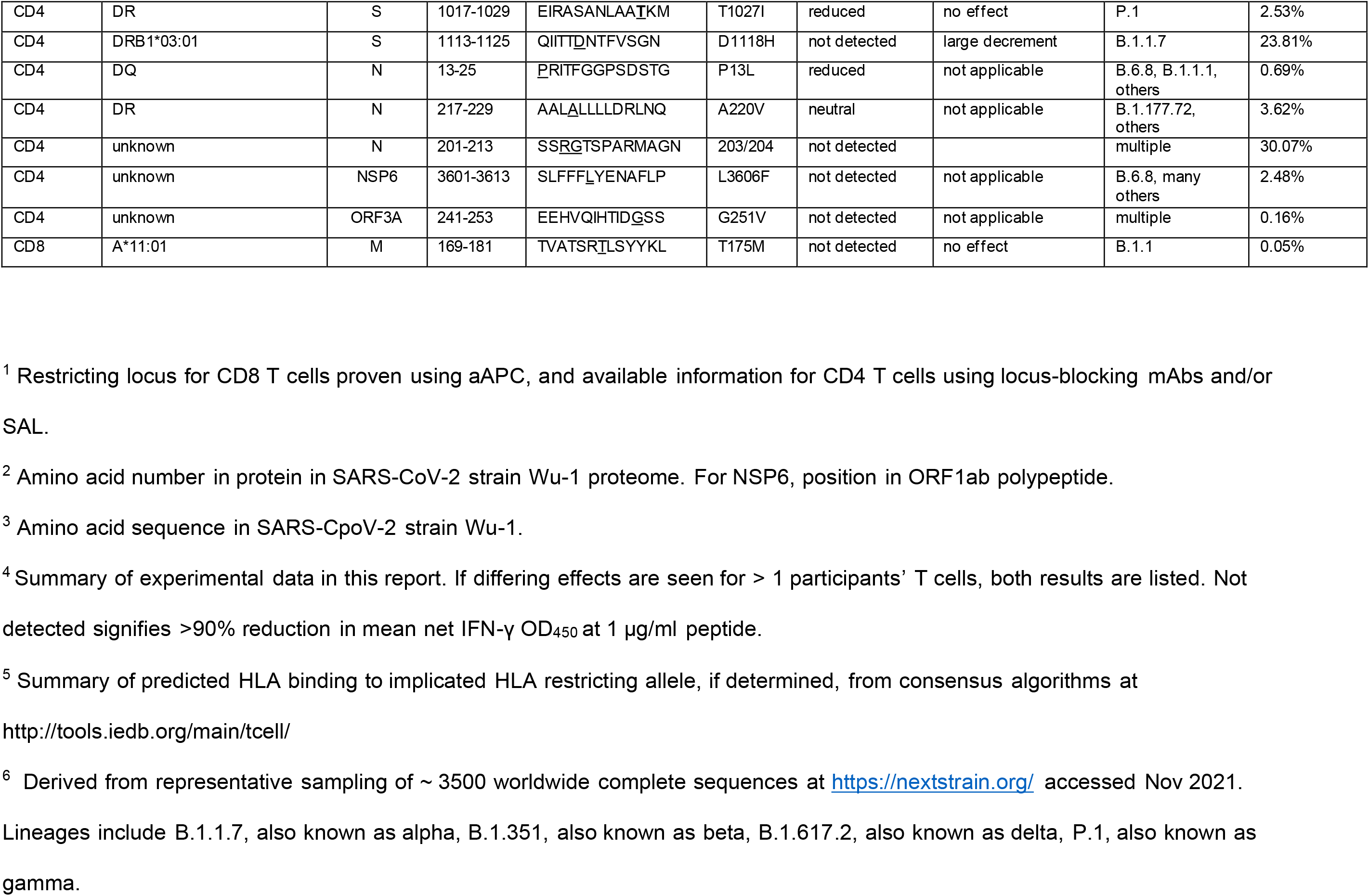
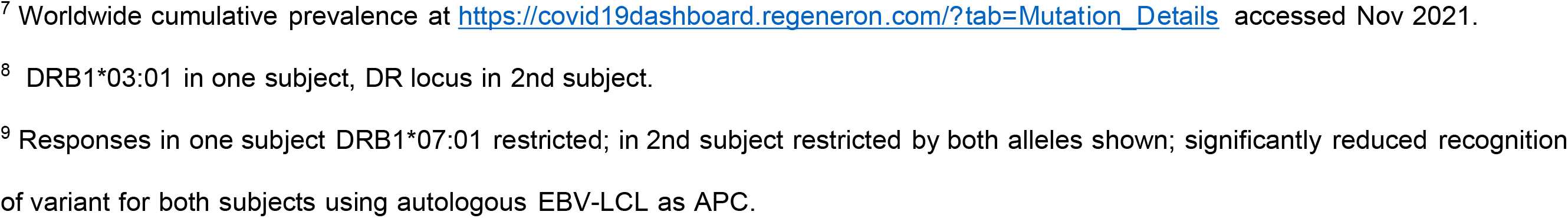
Summary of varied T cell recognition of SARS-CoV-2 peptides from strain Wu-1 and variants.

Variant changes could also influence CD4 T cell recognition (primary data, Supplementary Figs. 14-17). In addition to loss of recognition of variant aa in B.1.1.7 (effects summarized in Table 1 and Fig. 7) and/or P.1 lineages, we also noted strain-specific recognition of B.1.1.7-associated sequences that were already prevalent in early 2020 and not unique to VBM (example, Supplementary Fig. 10, indicated in blue). A graphical summary of variants evaluated in S (Fig. 7) shows a mixture of responses that are either preserved, partially reduced in dose-response assays, or abrogated when tested with variant peptides.

## Discussion

The T cell response to SARS-CoV-2 is functionally important (31). Studies have correlated T cell kinetics, magnitude or phenotypes with disease in apparently immune competent persons with delayed or dis-coordinated immunity and poor clinical outcome (8, 32). T cell responses have been linked to protection from re-infection (33). Disease states and iatrogenic treatments that decrease T cell responses can prolong live virus shedding (34, 35). Cooperation between T and B cells, such as antigen-specific CD4 T_FH_ (36), also suggest that T cells are involved in pathogen control. Supporting this, S-specific CD4 T cell TCR breadth and depth correlate with nAb titers in COVID-19 convalescent persons (9). CD8 T cell depletion data in non-human primates indicate that these effectors contribute to vaccine-induced protection (3).

Many groups have detected responses to SARS-CoV-2 epitopes using peptide-based technologies (31), but less work has focused on T cell reactivity to complex antigens such as whole virus or full-length proteins. Both T cell priming, and effector responses of memory T cells, occur *in vivo* in the context of viral infection and/or loading of antigen into various APC, with important differences for HLA class I and II (37). In the present report, we leveraged the ability of moDC to cross-present whole viral antigen to CD8 T cells, and to present viral antigen to CD4 T cells, to enrich polyclonal SARS-CoV-2-specific T cells. Similar studies are rare; in one report, live virus induced subtle IFN-γ expression in convalescent T cells, but confirmation of virus specificity was not documented (38). Using established (25, 30, 39–42) readout methods, TCL created after whole virus or simpler antigen stimulation were used to study the breadth and specificity of the T cell response, reactivity to virus-infected cells, and recognition of sCoV and SARS-CoV-2 variants.

Acute COVID-19 most severely effects the respiratory tract, and while autopsy surveys can detect virus body-wide (43), it is likely that T cell contributions to host defense and possibly to inflammatory damage occur mostly in the respiratory tract and draining lymph nodes. Cytotoxic T cells have the potential to kill SARS-CoV-2 infected cells and thus reduce viral progeny and further cell infection. To date, reports of the ability of T cells to recognize SARS-CoV-2 infected cells are limited. We have begun such studies using the HBEC3-KT-A. We previously showed that HBEC3-KT-A cells are permissive for expression of SARS-CoV-2 ORF6 protein after infection, (44) and that infected cells stain positively for S protein and viral RNA (45). We find that HLA-matched S-specific CD8 T cells recognize virally infected cells. HBEC3-KT cells can differentiate into ciliated and mucus-producing goblet cells *in vitro* (46), and *in vivo*, ciliated respiratory epithelial cells have the highest levels of SARS-CoV-2 RNA (47). Zhang *et al.* have reported that the SARS-CoV-2 ORF8 protein can down-regulate HLA class I and limit CD8 T cell killing of peptide-sensitized target cells (48). Wagner *et al.* have shown recognition of SARS-CoV-2-infected cancer-origin lung alveolar cells by overexpressing ACE2 and exogenous HLA class I alleles. Effector cells were allogeneic T cells engineered to express TCRs specific for SARs-CoV-2 (49). Many viruses encode CD8 T cell evasion functions, effects that can be selective for non-transformed, physiologically relevant APC (50). We are currently optimizing SARS-CoV-2 infection of primary human nasal epithelial cells, differentiated to ciliated and goblet phenotypes, and pursuing CD8 recognition studies of other SARS-CoV-2 proteins, target cell killing, and reduction of viral yield in CD8 T cell studies.

A cytotoxic CD4 T cell phenotype has been detected during acute SARS-CoV-2 infection (51, 52). Upper and lower respiratory tract epithelial cells can display HLA class II required for CD4 T cell recognition, and pulmonary alveolar macrophages are excellent APC for CD4 T cells and are abundant in COVID-19 pneumonia, containing SARS-CoV-2 antigen (53). Thus, CD4 T cell recognition of whole viral antigen could be relevant to lung infection, again with either salutary or damaging effects.

We studied T cell responses from relatively few persons and have not attempted to evaluate novelty per-epitope given the plethora of work in this area. Coverage of the entire viral proteome may, however, enable some novel insights. Mass spectroscopy suggests a large spectrum in SARS-CoV-2 NSP abundance in infected cell cultures (54, 55) and *in vivo* (56, 57). Previous studies suggesting that viral protein abundance is associated with CD4 T cell immunodominance (39, 58) are supported by our finding of prevalent and poly-specific responses to the abundant S and N proteins. The high population prevalence of CD4 responses to M, NSP3 and ORF3A also agree with prior work (31). We also found responses to NSP2 and ORF9B in at least half the subjects studied proteome-wide. NSP2 has previously been described as a CD4 immunogen in convalescent persons, while ORF9B was not included in some previous surveys (4, 11, 59–62). ORF9B is an alternative open reading frame within the nucleoprotein locus encoding a 98 aa-long ORF9B protein that is abundant in infected cells (54), localizes to mitochondria (63), elicits strong antibody responses (64), and is involved in evasion of type I IFN responses (65). CD4 T cell breadth was studied in 17 samples from 8 persons. We found a median CD4 breadth of 7 viral antigens/specimen, and that 6 of 8 persons recognized 7 or more proteins. In contrast, a survey of 99 convalescent persons with proteome-wide peptides and an *ex vivo* AIM approach found an average of 3.2 CD4 T cell antigenic proteins per person with no subject recognizing over 6 proteins (4).

TCL allowed tests of a large number of SARS-CoV-2 variant peptides. We found many examples of abrogation or reduction of recognition of variants, some from variants of concern. The literature consensus is that T cell responses to infection or vaccination are poly-specific, such that failed variant epitope recognition may not have a net deleterious effect on immunity (31). However, detailed investigations remain warranted. Tarke *et al.* tested PBMC from convalescent persons infected early in the pandemic (66). While *ex vivo* T cell responses in convalescent PBMC measured by AIM did not differ between ancestral- and variant-derived peptide pools for CD4 or CD8 T cells, IFN-γ ELISPOT detected decreased overall for VOC compared to Wu-1 peptides. Agerer *et al.* focused the HLA A*02:01-restricted response to the S aa 269-277 YLQ epitope (67). The L270F variant showed decreased binding to HLA A*02:01 and no activation of YLQ-specific CD8 T cells. L270F has been detected in 3 of > 1.4 million global sequences. In contrast, the P272L variant we investigated has been noted over 9000 times. aa 272 is modeled to face TCR (68) and to not reduce HLA A*02:01 binding. Based on structural models (68) and other epitope variants (69, 70), the S P272L (epitope position 4) and K417N or K417T (epitope position 1) variants studied herein could generate variant-specific T cells, a hypothesis testable in convalescent PBMC. Recently, de Silva et al. documented additional examples of strain-specific CD8 T cell recognition of SARS-CoV-2 variants (71). COVID-19 vaccines mostly use Wu-1-like S antigen, such that vaccinee T cells may also detect variant-specific T cell recognition. Recently, Keeton et al. detected modest differences in CD8 T cell recognition of S between vaccine strain and delta variant peptides (72). In another vaccine study (66), AIM responses to the B.1.351 S pool were lower for CD4 and CD8 T-cells compared to Wu-1 peptides. The precise epitope and HLA restriction information in this report can focus studies of variant recognition in cases of breakthrough infection as the pandemic continues to evolve, and as epitope-based (NCT04776317) and variant-based vaccines (73) are studied in clinical trials.

The present report also adds to the growing body of literature concerning cross-recognition of sCoV and SARS-CoV-2 (20, 27, 74–76). The use of whole viral antigens in both stimulation and readout stages of our investigations of sCoV allowed us to document that polyclonal CD4 TCL from COVID-19 convalescent persons can recognize whole OC43 and 229E viral antigen. As mentioned above, this assay format integrates antigen processing and presentation to supplement studies that focus solely on peptide reactivity. Validation to the antigen, peptide, and HLA restriction level was obtained for an epitope in OC43 S. The presence and sequencing of SARS-CoV-2 and sCoV infections in our study population are unknown. However, the timing of our specimen collection shortly after recovery from COVID-19, and sero-epidemiologic data that most adults have been infected with multiple sCoV, suggest the T cell cross-reactivities we detected may reflect prior sCoV infections boosted by SARS-CoV-2, Data concerning sCoV T cell cross-reactivity can be integrated with antibody-based (77, 78) and clinical studies (79) to determine if sCoV-specific immune memory modulate COVID-19 disease, and to guide development of possible pan-coronavirus vaccines.

The current study has limitations. We observed no AIM response to whole SARS-CoV-2 in HD, yet, several groups have documented the presence of SARS-CoV-2-reactive T cells in uninfected individuals from either the naïve or sCoV-primed memory repertoires (20, 80). Our approaches may be better suited to memory response to SARS-CoV-2 infection itself but could be adapted to capture these responses, for example by using higher input cell numbers. We noted only moderate preservation of TCR sequences comparing *ex vivo* sorted AIM (+) T cells and expanded cultures. Biological factors related to TCR repertoire differences could include rare clonotypes that assorted into either the *ex vivo* TCRseq or expansion fractions of the AIM (+) cells, rather than being represented in both. Indeed, sequencing of AIM (+) cells ex vivo shows broad and diverse responses at the TCR sequence level even to single epitopes, with many clonotypes detected at low levels (81). T cell programming could render some T cells refractory to expansion, with an exhausted T cell phenotype noted for SARS-CoV-2-specific memory T cells in some settings (82). Technical factors could also contribute, ranging from DNA extraction or PCR inefficiencies during TCR sequencing protocols to lack of provision of T cell expansion culture conditions required for some of the AIM (+) cells. Analytically, we discounted T cell clonotypes detected at less than two copies, potentially reducing concurrency between *ex vivo* and expanded repertoires. Alternative methods such as single-cell TCRseq (83) to capture the TCR repertoire AIM (+) cells *ex vivo* can be applied in the future to study the virus-specific repertoire before and after expansion and to determine if there are transcriptional programs correlated with failure to expand. In addition, a relatively low number of persons were studied. The viral antigens used could have low abundance for some viral proteins in our antigen preparation, leading to underestimation of antigen breadth.

In conclusion, T cells are a functionally important component of the specific acquired immune response to SARS-CoV-2 infection and vaccination. We have shown brisk recognition of whole SARS-CoV-2 by both CD4 and CD8 T cells from convalescent persons, provide estimates of the integrated frequency of these cells in the circulation, and used proteome-covering tools to approach within-subject antigenic breadth at the antigen and epitope levels. We found many examples of loss of recognition of epitope variants by effector T cells recovered from persons infected early in the pandemic, and used whole viral antigens to document coronavirus cross-reactive T cell immunity.

## Methods

### Subjects and specimens

The COVID-19 cohort has been described (7–10). Subjects reporting SARS-CoV-2 detection and an illness compatible with COVID-19 were recruited. Subjects were seropositive for serum IgG for SARS-CoV-2 S and nucleoprotein (N) (7) (Supplementary Table 1). PBMC were cryopreserved in liquid nitrogen. Adult healthy donor (HD) PBMC were collected before 2019. Subjects provided informed written consent and studies were Institutional Review Board-approved. HLA typing was by PCR amplicon sequencing at Scisco Genetics (Seattle, WA).

### Antigens

For whole viral antigen, SARS-CoV-2 strain Washington 1 (WA1) (84) (Genbank MN985325.1) was cultured on Vero-E6 USAMRIID cells (gift of Dr. Ralph Baric, University of North Carolina, Chapel Hill, NC) at MOI 0.1 for 48 hours to 80% cytopathic effect. Cells were recovered by scraping and frozen/thawed thrice. Titers ranged from 4.2 x 10^7^ to 4.8 x 10^8^ pfu/ml in Vero-E6-USAMRIID plaque assay (85) before inactivation. After UV light (900 mJ/cm^2^) and three cycles of freeze-thaw, plaque assays were negative on Vero-E6-USAMRIID. Vero mock antigen was prepared in parallel. Full-length SARS-CoV-2 codon-optimized molecular clones non-structural protein (NSP)1-NSP10, NSP12-NSP16, S, ORF3A, ORF3B, E, M, N, ORF6, ORF7A, ORF7B, ORF8, N, ORF9B, ORF9C (also known as ORF9Bwu and ORF14) from Wuhan-Hu-1 (Wu-1, Genbank NC_045512.2), and ORF10 from strain HKU-SZ-005b (Genbank MN975262.1) cloned into pDONR207 or pDONR223 (29) (ThermoFisher, Waltham, MA), were obtained from Addgene (Waterton, MA). Gateway^TM^ reactions (ThermoFisher) shuttled inserts to expression vectors pDEST103 and pDEST203 for CD8 and CD4 T cell research (30). pDEST103 expresses proteins intracellularly as fusions after N-terminal eGFP driven by a CMV promoter. pDEST203 expresses 6-histidine fusion proteins under the control of a T7 promoter and is used (30) to express antigens via *in vitro* transcription/translation (IVTT) (Expressway, ThermoFisher). SARS-CoV-2 S Wu-1 with the D614G mutation, HDM_Spikedelta21_D614G (Addgene #158762), and isogenic mutant K417N are described (86, 87). Additional plasmids were designed per S consensus data (22) (88) concordant with representative sequences from GISAID (89) (EPI_ISL_760400 (B.1.1.7 lineage) and EPI_ISL_700420 (B.1.351 lineage)). These were designed with the same 21 aa C-terminal deletion as Wu-1 D614G and D614G/K417N and synthesized and cloned by Twist (South San Francisco, CA) into pHDM to create HDM_Spikedelta21_B.1.351 and HDM_Spikedelta21_B.1.1.7 for transient transfection of eukaryotic cells. The predicted polypeptide sequences of strains WA1 used for PBMC stimulation, Wu-1 used for most peptides differ at ORF8 aa 84 with an S residue in WA1 and L in Wu-1.

sCoV OC43 (NR-52725), 229E (NR-52726), and NL63 (NR-470) were obtained from BEI Resources. OC43 was cultured on VERO E6 AT cells stably transduced with angiotensin converting enzyme 2 (ACE2) and transmembrane serine protease 2 (gift from Michael S. Diamond, Washington University, St. Louis, USA), 229E was cultured on Huh7 cells (90) and NL63 cultured on MK2-LLC cells (CCL-7, ATCC). Antigen was prepared by freeze-thaw and clarification (400 x g, 10 min, room temperature) of infected cells and UV-inactivated, with mock uninfected cell antigens prepared in parallel. Viral titers measured in the producer cells prior to UV were 1.6 x 10^5^ TCID_50_/ml (OC43), 2.3 x 10^6^ TCID_50_/ml (229E) and 2.0 x 10^2^ TCID_50_/ml (NL63). Antigens were tested at 1:40 (v/v) in T cell assays. S genes from OC43, 229E, NL63 and HKU1 (R619-M89-303, R619-M66-303, R619-M90-303, R619-M91-303; 166014, 166015, 166016, 166017, all Addgene) were subcloned into pDEST203 (91). IVTT-expressed antigens were tested at 1:1000.

Peptides covering the SARS-CoV-2 strain Wu-1 predicted proteome, 13 aa long overlapping by 9 aa, were obtained at ≥70% purity (Genscript, Piscataway, NJ) and dissolved at 20 mg/ml in DMSO (ThermoFisher). Peptides in SARS-CoV-2 ORFs polypeptides PP1a and PP1ab are numbered by positions in the polypeptide rather than within non-structural proteins (NSPs). Pools contained ≤ 54 peptides, maintaining 1 μg/ml final in-assay concentrations each, and ≤ 0.3% DMSO. Selected peptides containing variant aa, peptides shorter than 13 aa predicted to bind to HLA class I alleles, or peptides internal to antigenic peptides were also studied. Variants surveyed included variant 501Y.V1 lineage B.1.1.7 (alpha) (92), variant B501Y.V2 lineage B.1.351 (beta) (22), variant 501Y.V3 P.1 lineage, (gamma) (93), and lineage B.1.617.2, (delta) (nomenclature per US CDC Oct 2021 https://www.cdc.gov/coronavirus/2019-ncov/variants/variant-info.html#Interest). sCoV homologs for defined SARS-CoV-2 Wu-1 epitopes in S, N, and M proteins were determined by sequence alignment. If ≥ 1 sCoV homolog, amongst OC43, HKU1, 229E, and NL63, had identity at ≥ 7 aa with antigenic peptides from SARS-CoV-2 Wu-1, 13 aa homologs of the antigenic SARS-CoV-2 peptide from all 4 sCoV were assayed. In addition, OC43 S peptides, 15 aa long/11 aa overlap (EMPS-OC43-S-1, JPT, Berlin, Germany) were tested as pools or single peptides at 1 μg/ml final in ≤ 0.4% DMSO.

### SARS-CoV-2-specific T cell enrichment and expansion

For DC-AIM-based enrichment, monocytes were enriched (CD14 positive, Stemcell) and cultured in serum-free AIM-V (Stemcell) with 10 ng/ml each IL-4 and GM-CSF (R&D, Minneapolis, MN) in 6-well plates. Monocyte-origin DC (moDC) were collected at day 7 with EDTA/scraping, and seeded at 2.5 x 10^5^/well in 48-well plates. SARS-CoV-2 antigen was loaded into moDC by adding 25 μl UV- killed virus or mock antigen in 1 ml T cell medium (TCM). After 4 hours, 2 x 10^6^ autologous total T cells (negative selection, Stemcell) were added for 18 hours. Cells were stained with anti- CD3-PE (BioLegend, UCHT1), anti-CD4-APC-Cy7 (Becton Dickinson, RPA-T4), anti-CD8-FITC (ThermoFisher 3B5), anti-CD69-BV421 or PE-Cy7 (BioLegend, FN50), anti-CD137-APC (Becton Dickinson, 4B4-1), and 7-actinomycin D (7-AAD). Live CD3^+^CD4^+^CD8^-^ or CD4^-^CD8^+^ cells with CD137 and CD69 expression were bulk-sorted and 20% of sorted cells were non-specifically expanded for two cycles (94), with 80% saved for TCR sequencing. moDC were also used to present single viral proteins. HeLa cells (CCL-2, ATCC) were transfected with SARS-CoV-2 genes cloned into pDEST103, or pDEST103 control, using FuGene 6 (Promega) and collected at 48 hours. Transfected cells were suspended to 1 x 10^6^ cells/ml and UV-C-treated in a 6-well plate with 3600 μJ/cm^2^. moDC were collected at day 7 with EDTA and 1 x 10^5^ moDC in 0.4 ml TCM were seeded into 48-well plates for 4 hours prior to adding 1 x 10^5^ UV- treated HeLa for one hour. 1.5 x 10^6^ autologous PBMC in 0.1 ml were added at 37°C for 18 hours. AIM sorting was also used after stimulation of PBMC without moDC addition. For whole virus, UV-inactivated SARS-CoV-2 was incubated at 1:20 to 1:40 dilution with 1.5 x 10^6^ PBMC in 0.2 ml TCM in 96 well U-bottom plates for 18 hours. Mock virus negative control and 1.6 μg/ml phytohemagglutinin (PHA)-P (Remel, Lenexa, KS) positive control were included. For viral proteins, SARS-CoV-2 individual ORFs in pDEST103 were transfected into and expressed by COS-7 cells (#CRL-1651, ATCC, Manassas, VA), which were lysed with three cycles of freeze-thaw treatment. Pooled lysates representing multiple SARS-CoV-2 proteins were added to PBMC at a 1:1 volume ratio for 18 hours.

For proliferation-based enrichment, PBMC were labeled with cell-trace violet (CTV) (ThermoFisher) and cultured at 4 x 10^6^/well in 24-well plates in 2 ml (TCM) (25) for 5 days with SARS-CoV-2 peptide pools (≤ 54 peptides/pool, 1 μg/ml final each) covering S (6 pools), M (one pool), N (two pools), or NSP6 and E proteins (2 pools). After staining with anti-CD3-PE, anti- CD4-APC (ThermoFisher, S3.5), anti-CD8-FITC, and 7-AAD for viability, live CD3^+^CTV^low^ cells with CD4^-^CD8^+^ or CD4^+^CD8^-^ phenotype were bulk-sorted (FACSARIA II, Becton Dickinson). Polyclonal CTV^low^ T cell lines were expanded twice polyclonally (94).

### Tetramer sorting

Monomeric HLA-β2M complexes with UV-labile peptide (BioLegend) were used for UV peptide exchange and tetramerized with streptavidin-APC or streptavidin-PE (Becton Dickinson) per the manufacturer. Bulk-expanded CD8 T cells (∼10^6^) were stained in 100 μl TCM with 2 μl tetramer for 30 minutes on ice, followed by anti-CD4-APC-H7 and anti- CD8α-FITC. After 7-AAD staining, live CD8^+^CD4^-^tetramer^high^ cells were sorted, expanded (94), and cryopreserved. CD8 T cells recognizing Merkel cell polyomavirus (MCPyV) T antigen (T-Ag) aa 32-40/HLA A*03:01 were tetramer-sorted from Merkel cell carcinoma tumor-infiltrating lymphocytes (28).

### T-cell functional assays

CD8 TCL reactivity to viral proteins was measured using COS-7 artificial antigen presenting cells (aAPC) co-transfected with subject-specific HLA class I cDNA and SARS-CoV-2 ORFs, or fragments of SARS-CoV-2 ORF1a/1ab (95). HLA A and B cDNA alleles were amplified by RT-PCR (94) or synthesized (Genscript), cloned into pcDNA3.1(-) (ThermoFisher) (96) and sequence-verified. COS-7 in 96-well flat plates were co-transfected with 100 ng/well each HLA cDNA and SARS-CoV-2-p103 plasmids, using FuGene6 (96). After 2 days, 1 x 10^5^ CD8 TCL/well were added for 24-48 hours into 200 μl TCM, and secreted IFN- gamma (IFN-γ) measured by ELISA (96). For CD8 T cell responses to peptides, aAPC expressed HLA cDNA only. At 2 days, 1-10 μg/ml viral peptide or DMSO control was added with responder TCL, or aAPC were peptide-pulsed for 1 hour at 37 °C, 5% CO2 and washed prior to adding TCL. Alternatively, peptide specificity assays used autologous EBV-lymphocyte continuous line (EBV-LCL) as APC at 2-5 x 10^4^ cells/well in 96-well U-bottom plates, in duplicate or triplicate, with 5-10 x 10^5^ CD8 TCL responders/well. Single or pooled peptides were added at 1 μg/ml each final in ≤ 0.3% DMSO. For a peptide to be listed as a CD8 T cell epitope, we required recognition (IFN-γ OD_450_ > 2X DMSO control) of 1 μg/ml or less peptide in ≥ 2 independent assays. To measure CD8 T cell recognition of SARS-CoV-2-infected cells, human bronchial epithelial cell 3 immortalized with cyclin dependent kinase 4 and human telomerase reverse transcriptase (HBEC3-KT) (97) transduced with ACE2 (44) (HBE3-KT-A) were infected at MOI 2 in 6-well plates using strain WA1-GFP (gift of Dr. Ralph Baric), or mock-infected, for 24 hours, harvested with Accutase (Thermo-Fisher) and plated at 20,000 cells/well in 96 well U-bottom plates. CD8 T cells were added (100,000/well) to a final volume of 200 μl TCM. Supernatants after 24 hours were assayed by IFN-γ ELISA (98).

CD4 T cell assays for whole virus and IVTT proteins used PBMC as APC. In duplicate or triplicate 96-well U-bottoms, 5-10 x 10^4^ each autologous PBMC and CD4 TCL were seeded in 200 μl TCM, with whole UV-treated SARS-CoV-2 or mock antigen (1:20-1:40), IVTT preparations (1:1000-1:2000) from SARS-CoV-2 or empty vector or HSV-2 gene-containing (99) negative controls, single or pooled peptides or DMSO control, or PHA-P (1.6 μg/ml) positive control. T cell activation was determined by supernatant IFN-γ by ELISA at 1-2 days or ^3^H thymidine incorporation (proliferation assay at day 3-day 4 (100). When proliferation was measured, autologous PBMC were irradiated (3,300 rad, X-ray source). The criteria for positivity were that in duplicate assays, both raw CPM values were at least twice the average of the negative control wells containing irrelevant HSV antigens and media.

Alternatively, responses to whole virus and IVTT proteins were measured by intracellular cytokine secretion (ICS). Polyclonal CD4 TCL were CTV-labeled (25). Autologous PBMC and CD4 TCL (2-5 x 10^5^ each) were co-incubated in 200 μl TCM in 96-well U-bottoms with antigens or controls at concentrations listed above. Co-stimulatory mAbs (anti-CD28 and anti-CD49d, BioLegend) and Brefeldin A were added (25) and cells analyzed after 16-18 hours. Cells were stained with Near-IR live/dead (ThermoFisher), lysed with 1X FACS lysing solution (Becton Dickinson), permeabilization with FACS Perm 2 (Becton Dickinson), and stained with anti-CD3- PE, anti-CD4-FITC (ThermoFisher, S3.5), and anti-CD8-PerCP5.5 (SK1), anti-IFN-γ-PE-Cy7, (B27, Becton Dickson) and anti-IL-2 (MQ1-17H12, Becton Dickinson). Data were acquired with FACSCANTO and cytokine expression quantified for live, CTV-positive CD3^+^CD4^+^CD8^-^ single cells (FlowJo 10.7.1, Becton Dickinson). For ICS-based proteome screens of CD4 TCL, two criteria were both required to score a protein positive. The ratio of the percent of IFN-γ and/or IL-2 positive CD4 T cells with SARS-CoV-2 antigen compared with pDEST203 empty vector-derived IVTT product, was > 2. For subjects W002 and W011 with high IFN-γ background for pDEST203 empty vector, the percent of double-positive IFN-γ/IL-2 cells, only, was used. Second, the difference in cytokine-positive cells between a SARS-CoV-2 protein and empty vector was > 1%.

CD4 T cell peptide analyses used autologous EBV-LCL as APC. TCL (2-5 x 10^4^/well) and EBV-LCL (5-20 X 10^3^/well) were co-plated in 96-well U plates in duplicate or triplicate in 200 μl TCM. Peptides at 1 μg/ml each final concentration were added as pools or singletons in ≤0.3% DMSO, or DMSO negative control. After 1 to 2 days, supernatant IFN-γ was measured by ELISA. For some assays with high IFN-γ ELISA OD_450_ values for negative control stimuli, ELISA was performed with diluted supernatants. In order for a peptide to be reported as an epitope, two or more wells tested with 1 μg/ml peptide were required to have an IFN-γ OD_450_ at least twice above DMSO background in at least two assays. For proliferation assays, peptides (1 μg/ml) were tested using autologous EBV-LCL (1 X 10^4^/well, irradiated 10,000 rad) and responder TCL (5 X 10^4^/well) in duplicate. For a peptide to be reported as an epitope, both replicates had a count per minute (CPM) value more than 2-fold the average CPM of DMSO negative controls, and, in follow-up triplicate screens, at least two of the three CPM values were more than 2-fold average CPM of negative controls. To enumerate CD4 T cell reactivities per- subject, each reactive peptide was counted as a separate epitope. If two variant SARS-CoV-2 peptides were reactive for the same person, this was counted as one reactivity. Reactivities detected in more than one workflow or PBMC timepoint per person are counted once. Responses to the same or variant peptides in multiple participants were analyzed both separately and after collapse to unique peptides.

We defined CD4 TCL HLA restriction as published (40). First, serial dilutions of peptide (1, 0.1, or 0.01 μg/ml) were tested in triplicate using autologous EBV-LCL as APC without or with mAbs that block HLA DR, DP, or DQ (100). To determine allele-level restriction, engineered APC expressing single HLA class II heterodimers were selected (40) to match the study subject. Negative controls were cells parental to the single antigen line (SAL). SAL assays typically used 1 μg/ml peptide or DMSO control. Some used SAL washed after a 1-hour pulse at 37 °C with peptides in titration. CD4 TCL were added and IFN-γ secretion measured by ELISA. To estimate functional avidit, we set an index value of 100% to the difference between the mean IFN-γ OD_450_ values of 1 μg/ml peptide and of DMSO. A peptide concentration was scored as positive if at least two of three triplicates at that concentration yielded net IFN-γ OD_450_ values (raw values minus the mean of DMSO) at least 30% of this index value. For selected peptide epitopes an alternative or expanded dilution series was used.

### TCR sequencing and analysis

AIM (+) PBMC were flash frozen after sorting in a minimal volume of TCM and DNA manually isolated with the Qiagen blood kit for small samples. DNA from culture-expanded AIM (+) cells was isolated by Adaptive. T cell receptor beta (*TRB)* complementarity determining region 3 (CDR3) repertoire sequencing was performed at Adaptive Biotechnologies (Seattle, WA) using the Immunoseq^TM^ TCRBv4b platform. Two replicate DNA aliquots were processed in parallel and only productive TRB CDR3 gene rearrangements present in at least one read in both replicates were reported. Analyses focused on functional CDR3 amino acid identity.

### Data availability

T cell epitopes have been uploaded to IEDB(102) with accession numbers 1000861 and 1000866. TCR datasets are in ImmuneACCESS (Adaptive).

### Approval

By the University of Washington Institutional Review Board as Study00004312.

### Statistics and variant abundance

Proportions of *ex vivo* AIM (+) cells were compared within-person between mock and stimulated conditions using Wilcoxon matched-pairs signed-ranks test (Instat 3.10, GraphPad, San Diego, CA). Proportions of *ex vivo* AIM (+) cells after stimulation were compared between clinical cohorts using Mann-Whitney test. P values are two-tailed. Correlation between IFN-γ and proliferation results used linear regression and default parameters (Prism 9.1.0, GraphPad). To estimate the global prevalence of SARS-CoV-2 amino acid variants, the worldwide SARS-CoV-2 sequence dataset hosted at GISAID (89) was accessed (101) using the mutation details routine, while lineages containing specific mutations were queried via Nextstrain (23), which accesses a representative subset of GISAID.

## Data Availability

T cell epitopes have been uploaded to IEDB with accession numbers 1000861 and 1000866. TCR datasets are in ImmuneACCESS (Adaptive).

## Acknowledgements

We thank the participants, the University of Washington Virology Research Clinic and Immunology Cell Analysis Facility, the Flow Core at Seattle Children’s Research Institute, Jesse Bloom, Katharine Crawford, Keara Malone, Andrea Loes, and Allison Greaney for SARS-CoV-2 plasmids, Ralph Baric for Vero-E6-USAMRIID cells and SARS-CoV-2-strain WA1-GFP, and Thomas Snyder and Adaptive Biotechnologies for Immunoseq^TM^ sequencing.

