## Supplemental figures 1-17 for "T cell response to intact SARS-CoV-2 includes coronavirus cross-reactive and variant-specific components"

Supplementary Fig. 1

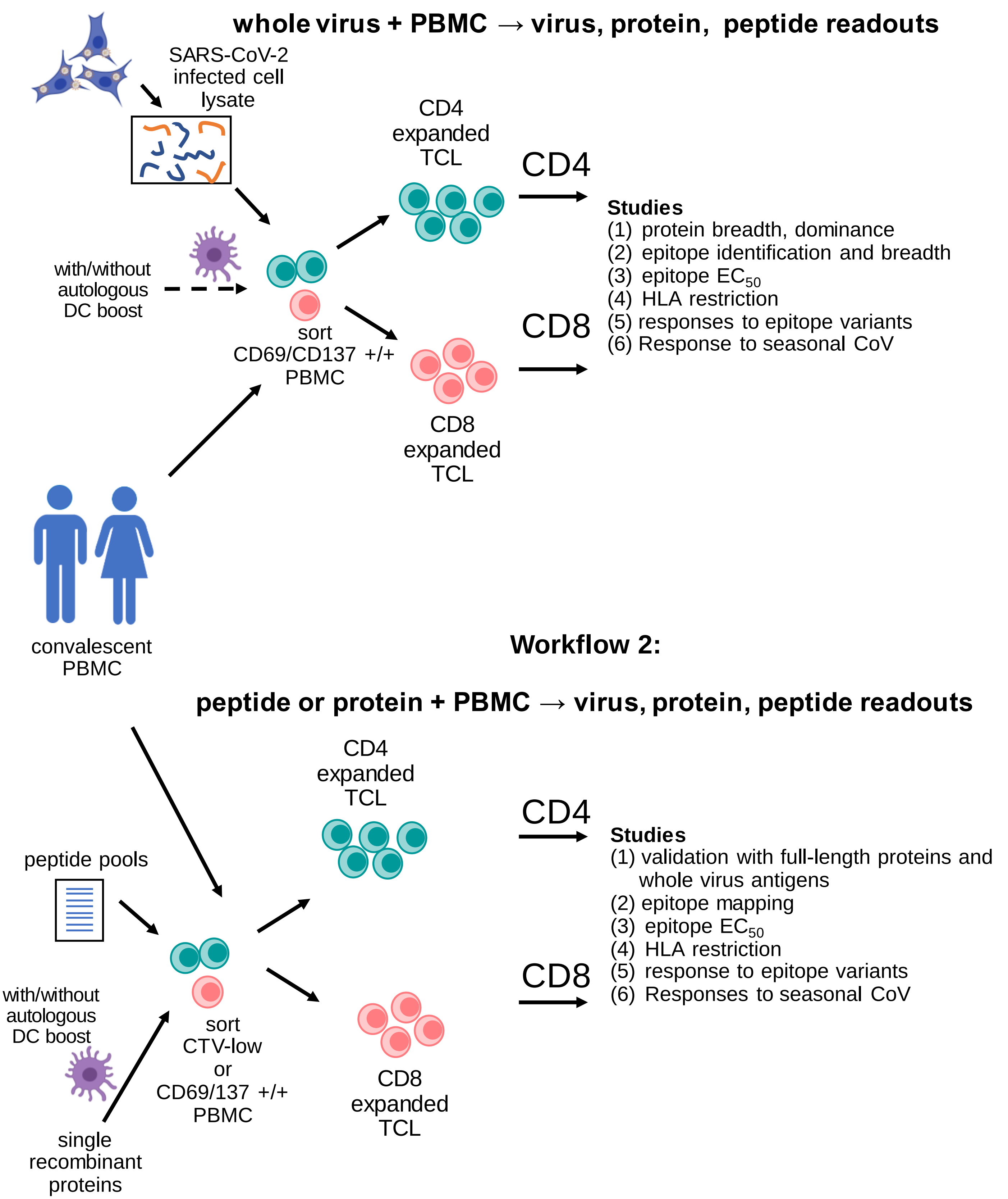


Supplementary Fig. 1. Schematic overview of SARS-CoV-2 T cell research workflows. Created with BioRender.com.

Supplementary Fig. 2

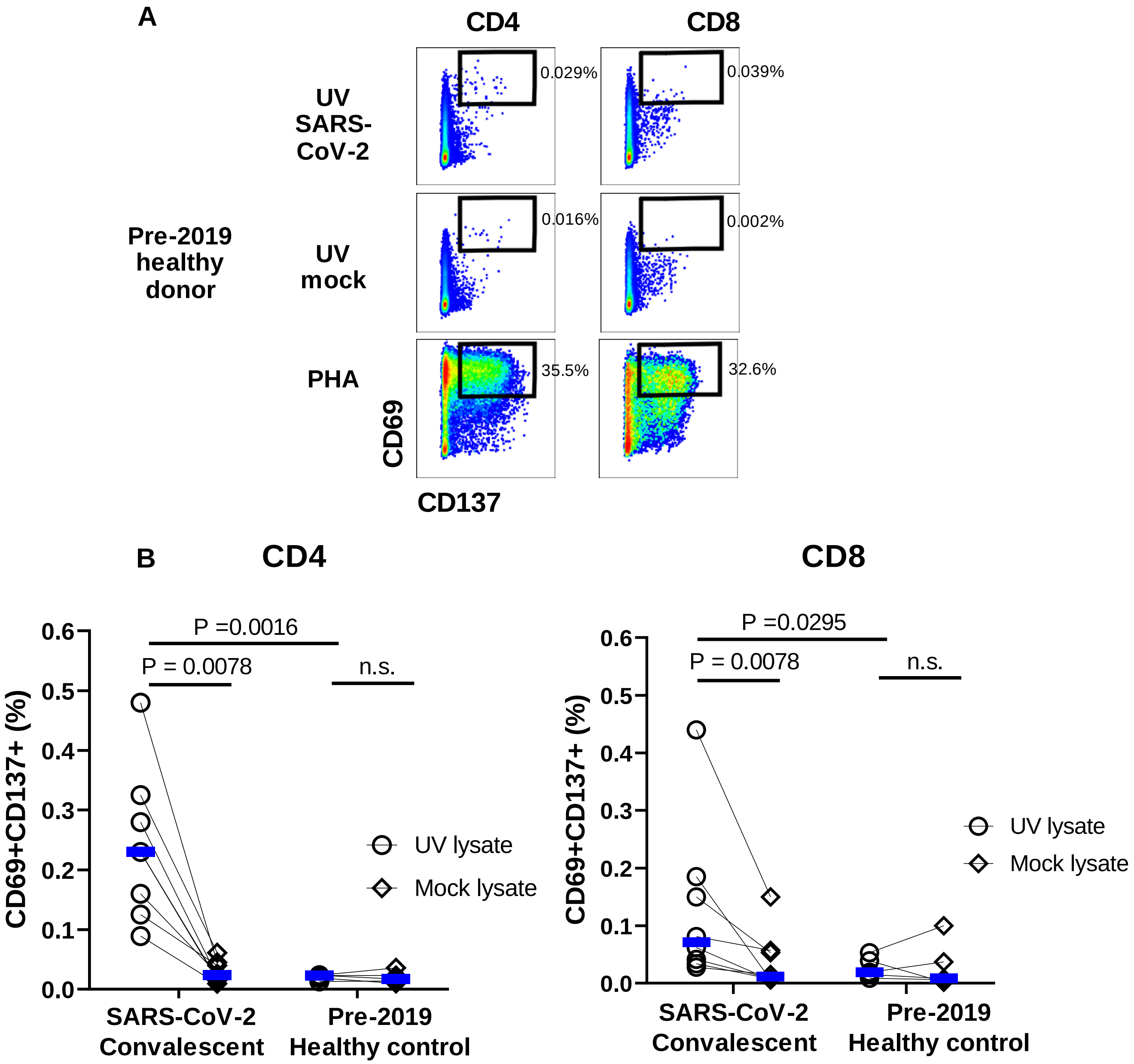


Supplementary Fig. 2. AIM assay distinguishes COVID-19 convalescent and control individuals. A: Representative pre-2019 healthy donor PBMC incubated 18 hours with the indicated stimuli. Gated T cells subsets are shown. Very low or absent net responses to SARS-CoV-2 are observed. Square gates indicate regions measured for percent AIM (+) cells. B: AIM (+) cell abundance in n=8 PBMC specimens from seven COVID-19 convalescent participants in response to whole killed SARS-CoV-2 or mock antigen. Dendritic cells were not used. There are net responses to viral antigen for convalescent specimens but not from n=5 pre-2019 healthy donor PBMC at right (Wilcoxon matched pairs signed rank test). Convalescent PBMC show greater AIM responses to viral antigen than to pre-2019 specimens (Mann-Whitney test). CD4 and CD8 T cell responses shown separately. Blue bars are median values. n.s. = not significant

Supplementary Fig. 3

A

B

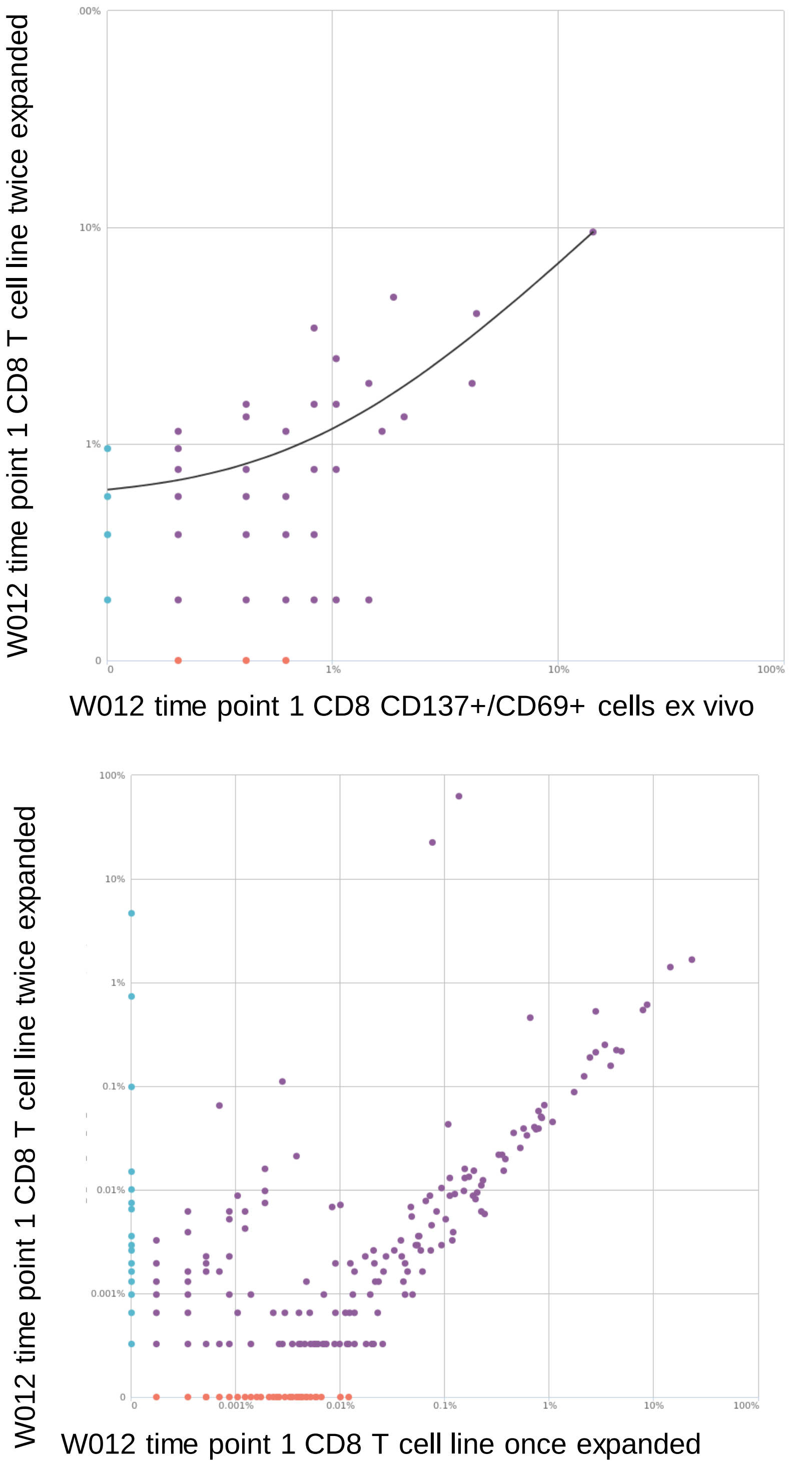


Supplementary Fig 3. A: Productive frequencies of T cell clonotypes from subject W012 at the TRB CDR3 AA level, comparing directly ex vivo sorted AIM (+) CD8 T cells with twice-expanded cells used for detailed studies. B: Similar analysis for the same Specimens after one or two culture expansions.

Supplementary Fig. 4

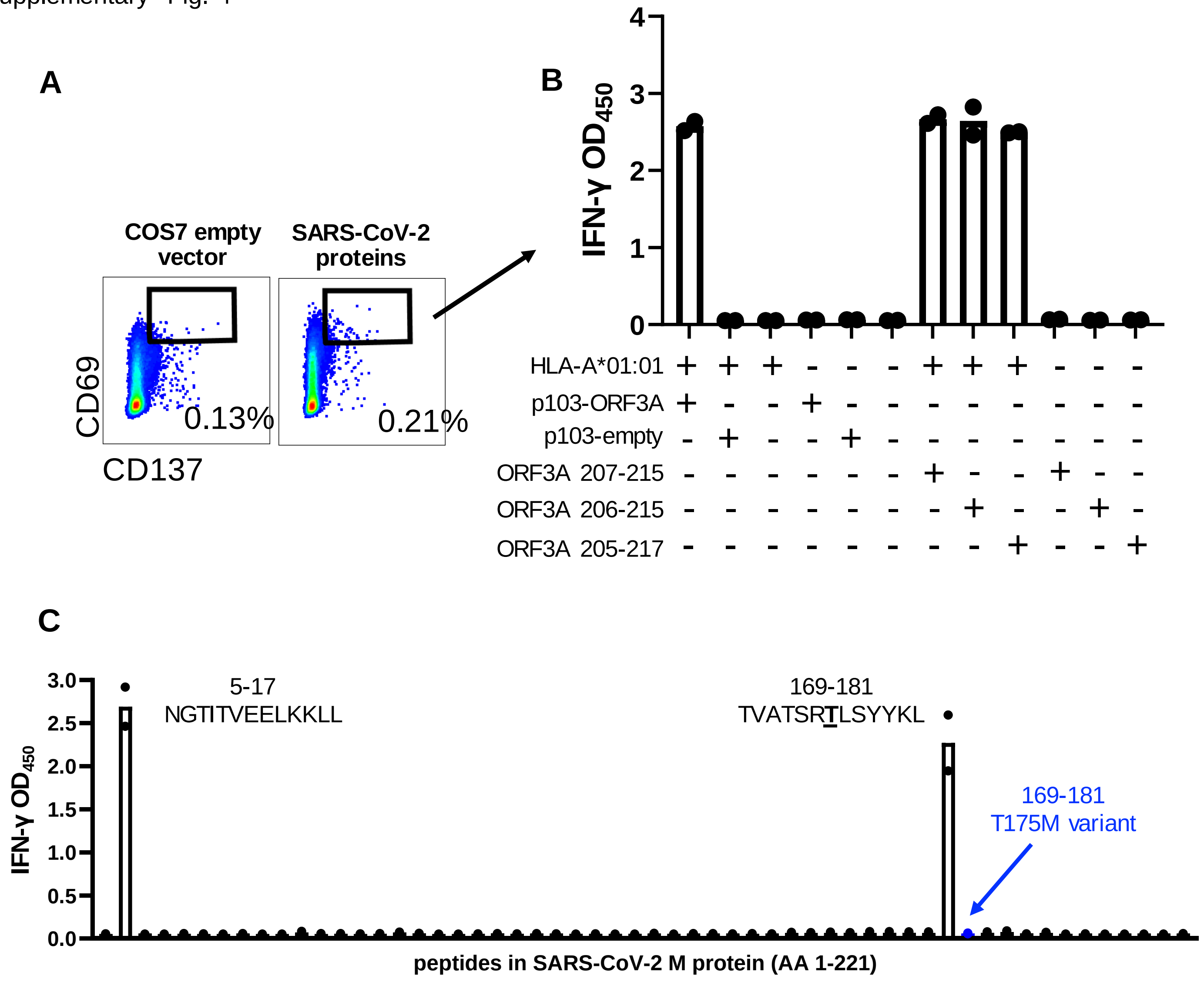


Supplementary Fig. 4. CD8 T cell epitopes in SARS-CoV-2 revealed by stimulating PBMC with simple antigens. A: PBMC were subject W006, specimen 3 were stimulated with DC and pooled SARS-CoV-2 proteins expressed by transient transfection of Cos-7 cells or empty vector negative control. Dotplots are gated live CD3+/CD4-/CD8+ cells. B: Cells in the indicated gate in A were sorted, expanded and tested against the indicated stimuli using Cos-7-based aAPC. Black dots are values for individual wells and bars are means. C: PBMC from subject W004 were stimulated with pooled peptides from M protein and proliferated cells were sorted and tested against individual peptides using HLA A*11:01 aAPC. The indicated 13 AA peptides were positive.

Supplementary Fig. 5

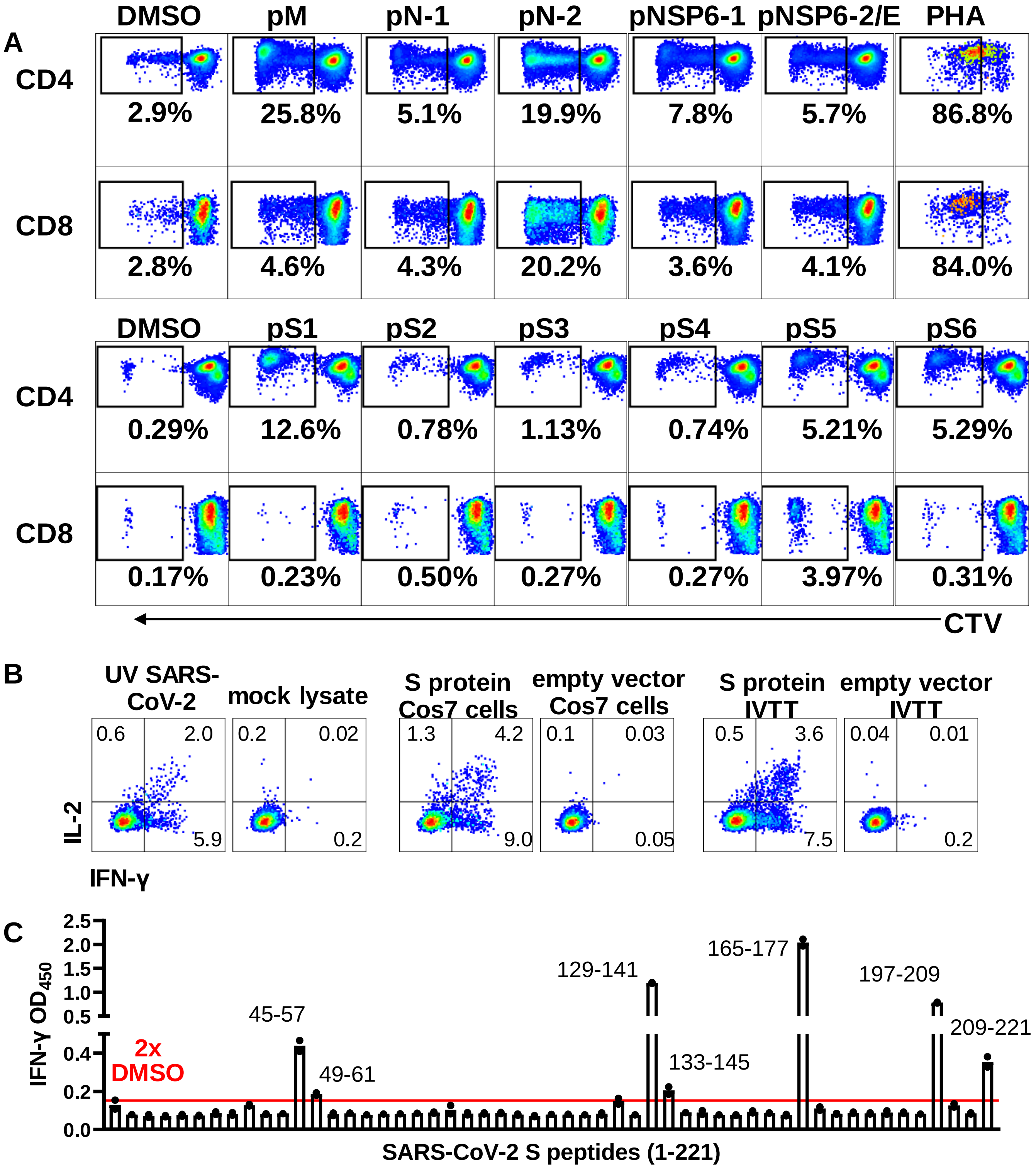


Supplementary Fig. 5. Proliferation-based isolation of SARS-CoV-2-reactive T cells. A: PBMC from subject W004 labeled with cell tracker violet (CTV) were incubated with DMSO or the indicated peptide pools covering SARS-CoV-2 strain Wu-1 M (membrane), N (nucleoprotein, 2 pools), or NSP6 (2 pools; E = envelope combined with NSP6 pool 2) in the top two rows, or DMSO and S (spike, 6 pools) in next two rows. Dotplots indicate percentages of CTV-low cells in gated CD4 (rows 1 and 3) or CD8 (rows 2 and 4) T cells. B: PBMC from subject W006, specimen 1, were stimulated with pooled SARS-CoV-2 S AA 1-221 peptides and CTV-low, CD4 T cells sorted and expanded. The resultant CD4 TCL responded to SARS-CoV-2 and full-length S protein expressed by Cos7 transfection or IVTT but not negative controls. Dotplots are gated live CD3+/CD4+/CD8- responder cells after de-gating of autologous PBMC used as APC. Numbers are percentages of cells in quadrants. C: The same TCL assayed against peptides in the pool used for stimulation. Autologous LCL were used as APC in duplicate assays. Amino acid numbers of peptide meeting criteria for positivity are displayed.

Supplementary Fig. 6

A

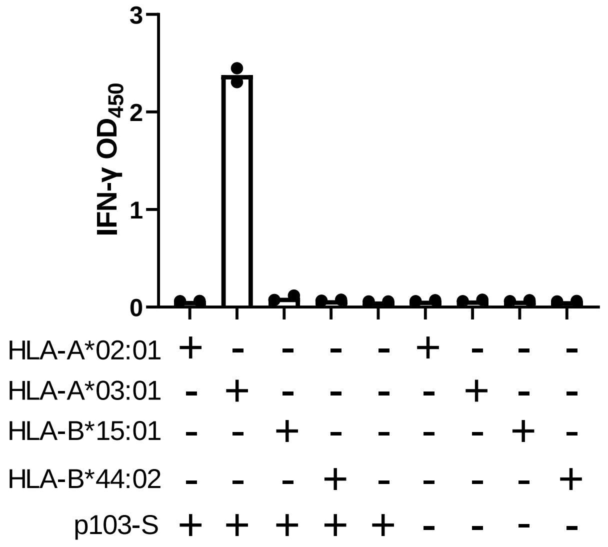


B

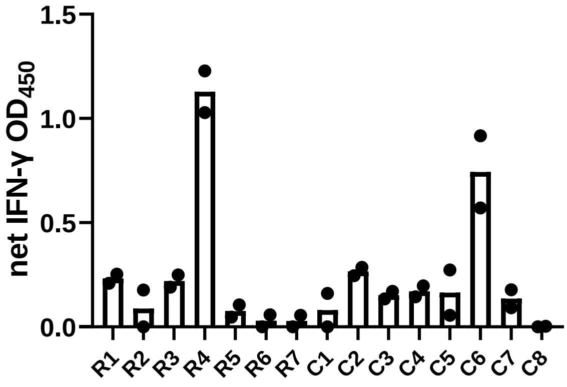


| | C1 | C2 | C3 | C4 | C5 | C6 | C7 | C8 |
|---|---|---|---|---|---|---|---|---|
| R1 | P1 | P2 | P3 | P4 | P5 | P6 | P7 | P8 |
| R2 | P13 | P14 | P15 | P16 | P17 | P18 | P19 | P20 |
| R3 | P25 | P26 | P27 | P28 | P29 | P30 | P31 | P32 |
| R4 | P37 | P38 | P39 | P40 | P41 | P42 | P43 | P44 |
| R5 | P49 | P50 | P51 | P52 | P53 | | | |
| R6 | P9 | P10 | P11 | P12 | P33 | P34 | P35 | P36 |
| R7 | P21 | P22 | P23 | P24 | P45 | P46 | P47 | P48 |

377-389 FKCYGVSPTKLND

C

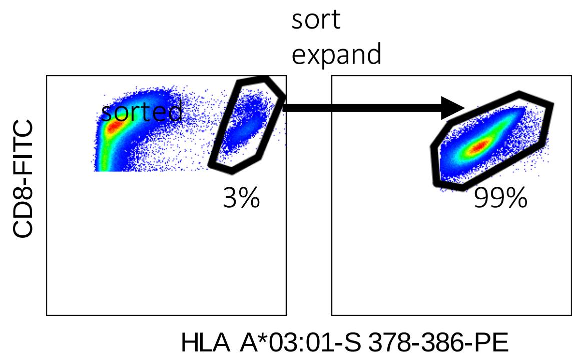


Supplementary Fig. 6. A: Reactivity of bulk CD8 TCL co-cultured with aAPC created by transfecting Cos-7 cells with the indicated plasmids. p103-S encodes SARS-CoV-2 spike. B: Mapping of reactive 13 AA peptide within SARS-CoV-2 S using row (R) and column (C) pools of overlapping peptides with reactive pools and intersection highlighted. The predicted peptide sequence is given. C: Detection of epitope-specific CD8 T cells with fluorescent tetramers of HLA A*03:01 complexed with S AA 378-386 before (left) and after (right) sorting and further cell expansion. Effector cells for these experiments were created by stimulating immune PBMC with autologous DC loaded with HeLa cells transiently transfected with S and sorted for CD69/CD137 expression followed by non-specific expansion.

Supplementary Fig. 7

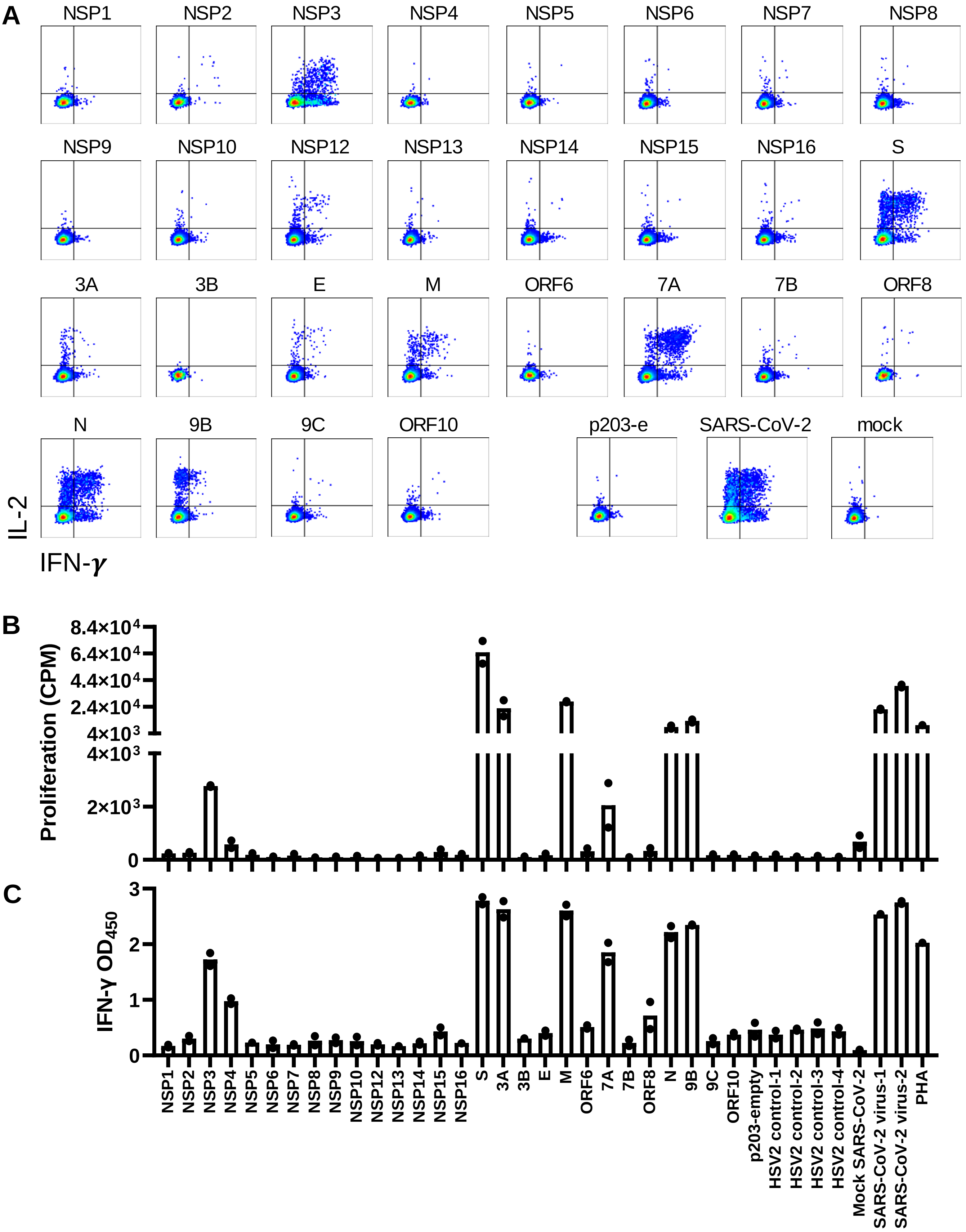

Supplementary Fig. 7. Proteome-wide CD4 T cell antigen determination for SARS-CoV-2. A: AIM-enriched, expanded CD4 T cells from subject W009 PBMC stimulated with whole virus, without DC, were tested by ICS against the indicated proteins. Positive control is whole SARS-CoV-2 antigen; negative controls are p203e empty vector protein and mock virus. Dotplots are gated for responder, live, single, CD3+/CD4+/CD8- T cells. B: AIM-enriched, expanded CD4 T cells from subject W001, time point 1, with use of moDC, were tested in duplicate proliferation (upper) and secreted IFN-γ ELISA (lower) formats. Data are individual values with bars representing averages. Positive controls are whole SARS-CoV-2 antigen and PHA; negative controls are p203 empty vector protein, random HSV-2 proteins expressed in p203, and mock SARS-CoV-2 antigen.

Supplementary Fig. 8

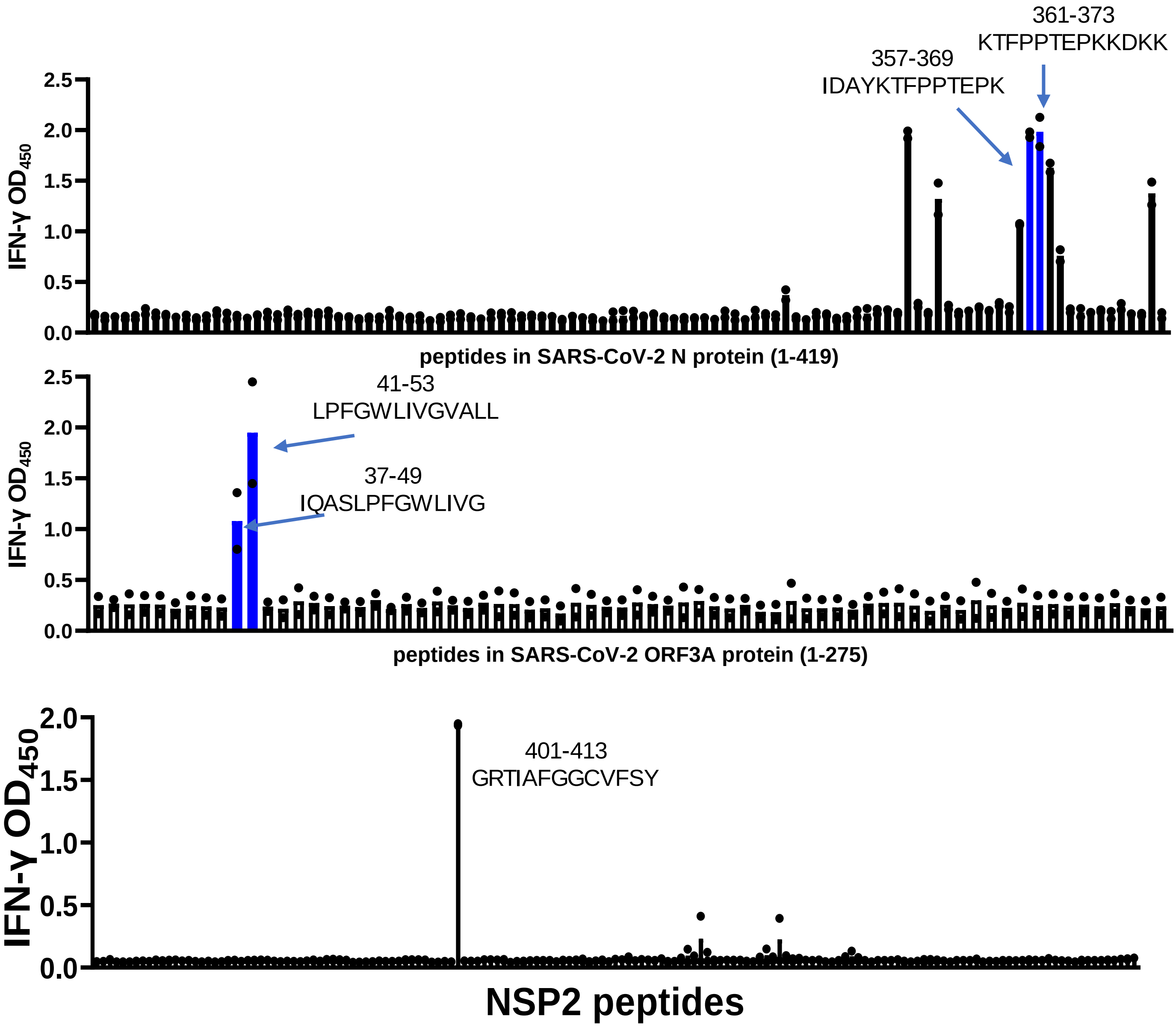


Supplementary Fig. 8. Peptide scans of SARS-CoV-2 N and ORF3A proteins for polyclonal CD8 TCL from subject W005 specimen 1, created using whole virus and moDC, using EBV-LCL used as APC. The peptides with blue bars in the top two screens and the NSP2 peptide were repeatedly reactive in follow-up assays and are reported as epitopes in this report. Black dots are values for individual wells and bars are means.

Supplementary Fig. 9

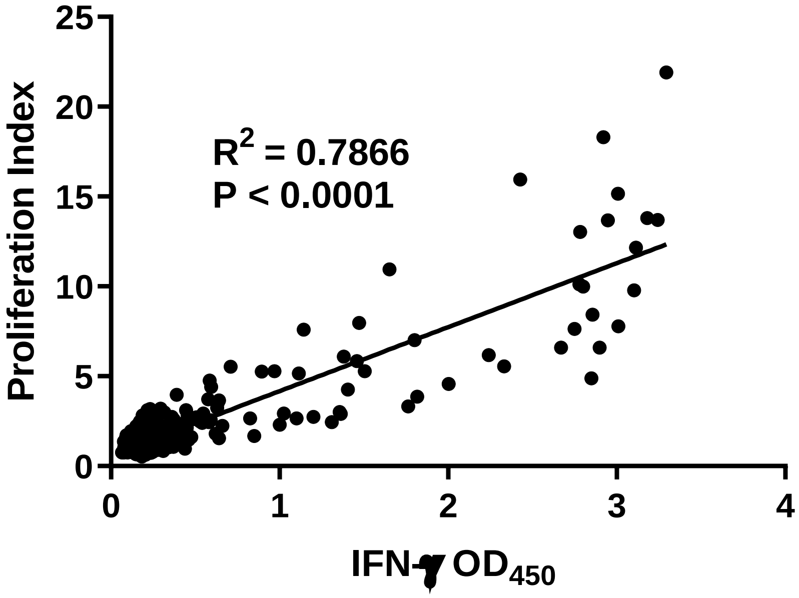


Supplementary Fig. 9. Correlation of T cell activation readouts. CD4 TCL from subject W001, specimen, 1 obtained by DC-boosted stimulation of PBMC with whole UV-killed SARS-CoV-2 and AIM sorting, was screened with peptides (n=319) covering SARS-CoV-2 S protein in duplicate. Each dot represents an assay well. The X axis is the IFN-γ ELISA $OD_{450}$. The Y axis is the proliferation index, defined as counts per minute (CPM) $^3$H thymidine incorporation divided by the average CPM from negative controls. Statistics are from linear curve fit.

Supplementary Fig. 10.

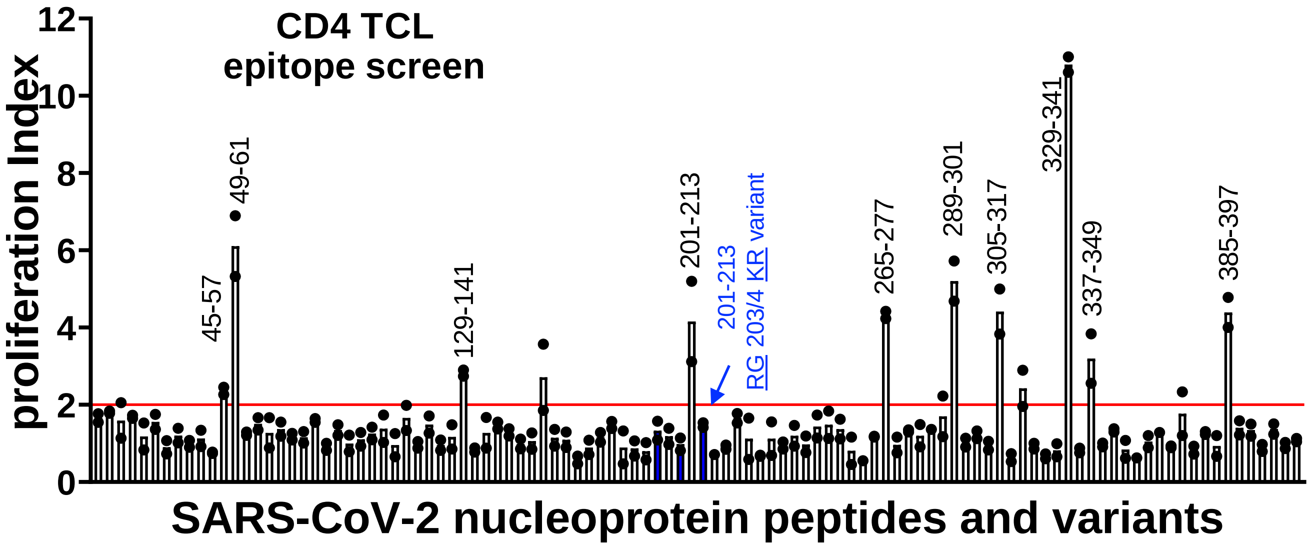


Supplementary Fig. 10. Epitope discovery using polyclonal SARS-CoV-2-reactive TCL originated using PBMC stimulation of DC loaded with whole viral antigen and AIM sorting. CD4 TCL from subject W001, specimen 1 tested in duplicate proliferation assay with irradiated EBV-LCL as APC and overlapping 13 AA nucleoprotein peptides. Peptides in blue have a substitution of RG for KR at nucleoprotein AA 203-204. Dots are proliferation indices of duplicate wells and bars are averages. Coordinates of peptides with duplicate proliferation indices each > 2 (red line) are labeled.

Supplementary Fig. 11

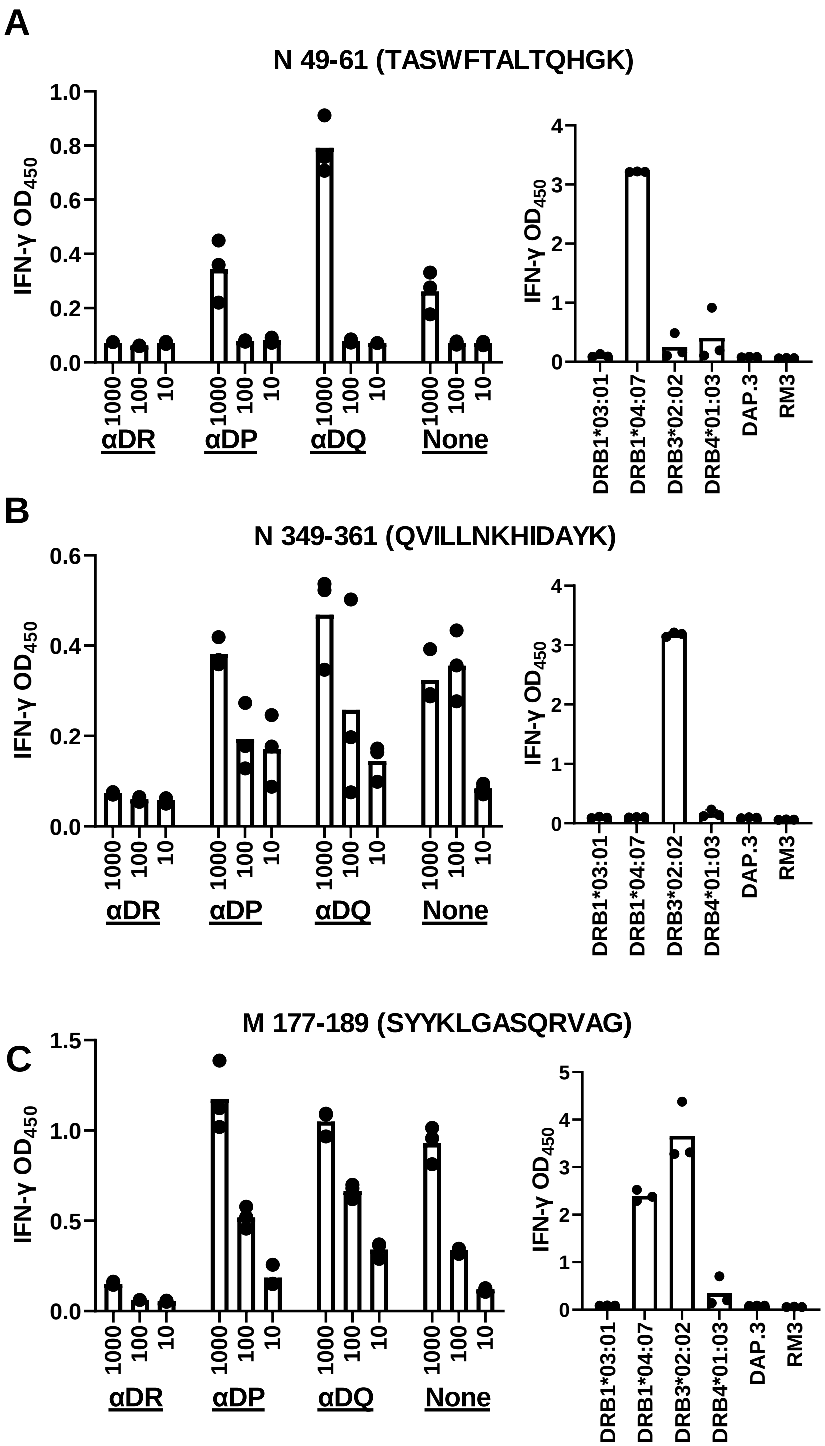


Supplementary Fig. 11. SARS-CoV-2 CD4 T cell epitope HLA restriction locus and allele definition. A: At left, CD4 TCL from subject W009 was incubated with serial dilutions of the indicated peptide from SARS-CoV-2 N protein 49-61 epitope in the presence or absence of locus-specific blocking mAb. At right, 1 μg/ml peptide was tested with the indicated aAPC. B: Similar workup for the same polyclonal CD4 CTL for a distinct peptide in N showing restriction by a different HLA DR allele. C: Dual recognition of M peptide 177-189 by both HLA DRB1*04:07 and DRB3*02:02. For each peptide, HLA-negative parental cell lines DAP.3 and RM3 are shown at right. Dots are data from triplicate assays and bars are mean values.

Supplementary Fig. 12.

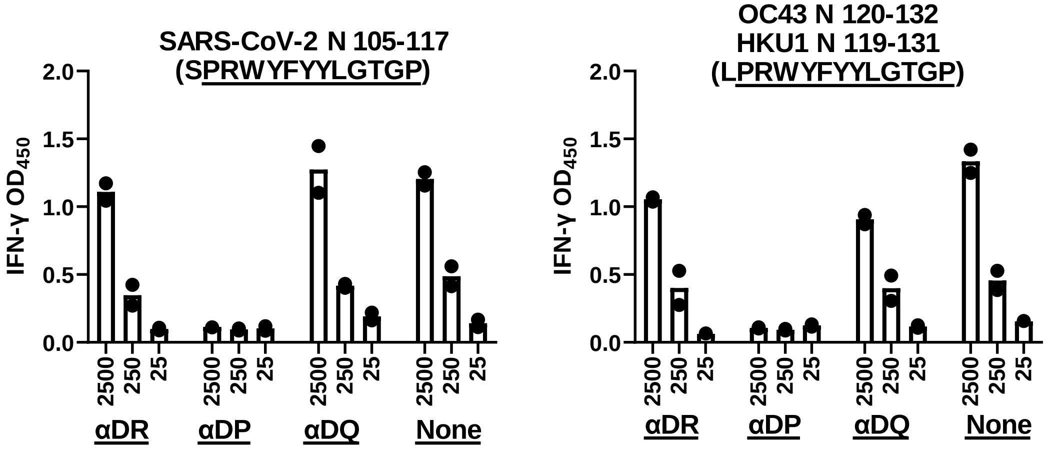


Supplementary Fig. 12. CD4 TCL from subject W005, re-stimulated form PBMC with whole SARS-CoV-2 antigen and moDC, recognize a near-identical peptide in SARS-CoV-2, OC43, and HKU1 N protein. The concentration of synthetic peptide testing in micrograms per ml is indicated on the X axis. All responses are restricted by HLA DP as indicated by inhibition with the indicated locus-specific mAb. Autologous LCL were used as APC.

Supplementary Fig. 13.

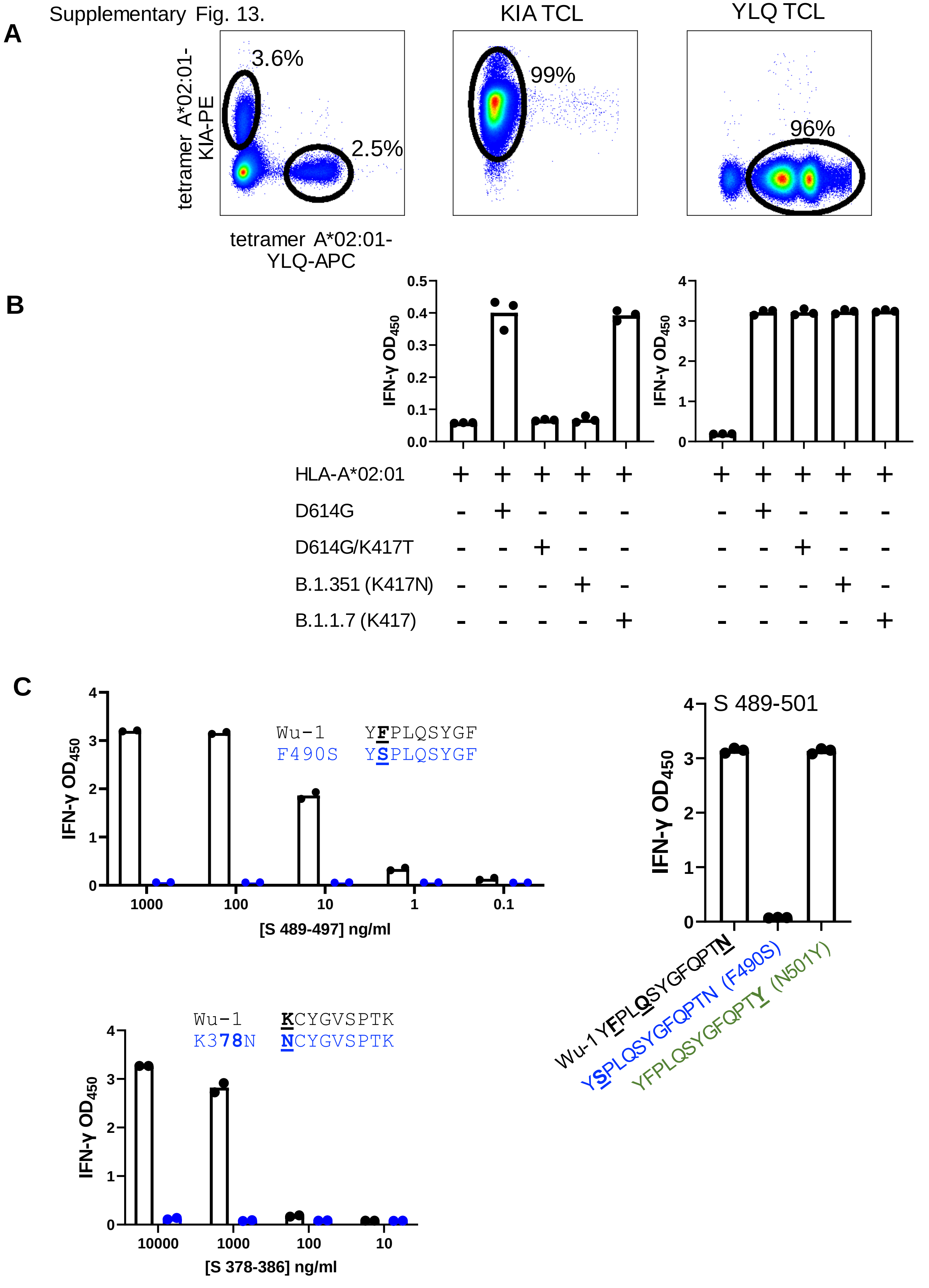

Supplementary Fig. 13. SARS-CoV-2 CD8 T cell recognition of variant peptides in Spike. A: Enrichment of spike YLQ-269-277 and KIA-417-425 CD8 TCLs with A*02:01 tetramers containing Wu-1 peptides from participant W004 cell line. Right panels show purity of expanded tetramer-positive cells. B: SARS-CoV-2 spike D614G (near wild-type) or variant plasmids were co-transfected with HLA-A*02:01 into aAPC. KIA-specific CD8 T cells failed to recognize spike mutants altered at AA 417. As controls, KIA-specific CD8 T cells recognize the B.1.1.7 spike plasmid with K417, and all spike variants are recognized by YLQ-specific CD8 T cells. C: Each panel shows the peptides tested from strain Wu-1 and selected variants. Data show peptide titrations except for spike 489-501 variants tested at 1 μg/ml only. In each case, Cos-7 transfected with the relevant HLA cDNA were used as aAPC. Dots are raw data and bars are averages of duplicate or triplicate assays. Data are from subject W008 (spike F490 epitope), and from subject W003 (spike K378 epitope).

Supplementary Fig. 14

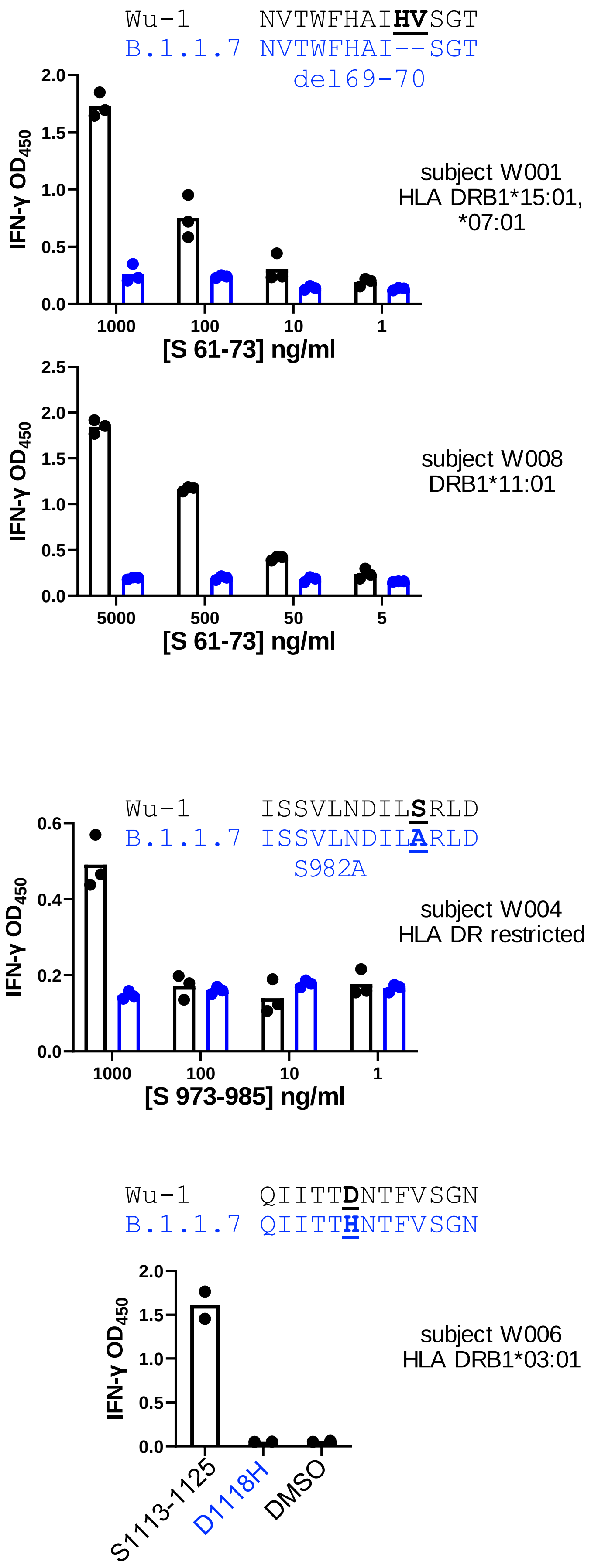

Supplementary Fig. 15

```
Wu-1     INITRFQTLLALH
B.1.351  INITRFQT---LH  242-244del
```

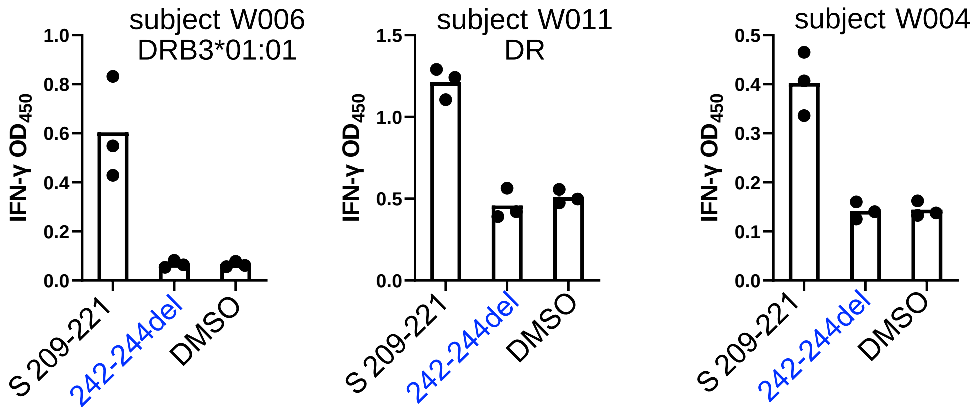


```
Wu-1     FQFCNDPFLGVYY         B.1.351  FQFCNYPFLGVYY  D138Y
B.1.617  FQFCNDPFLDVYY  G142D  B.1.1.7  FQFCNDPFLGV-Y  Y144del
```

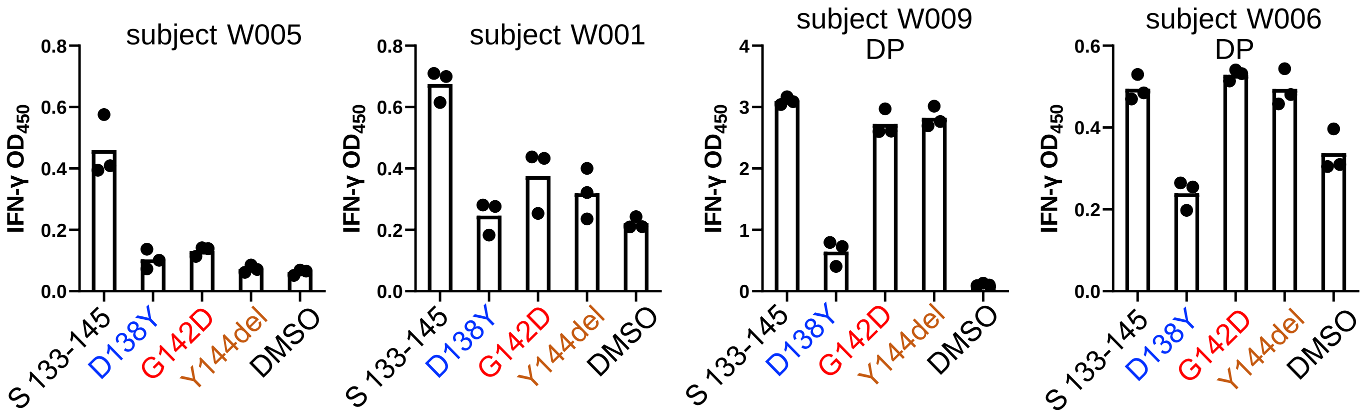


```
Wu-1     IAYTMSLGAENSV
B.1.351  IAYTMSLGVENSV  A701V
```

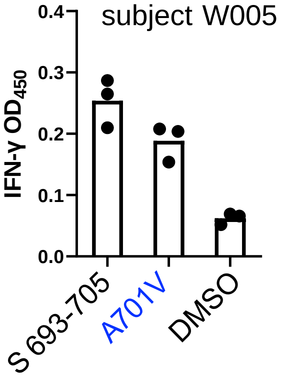


```
Wu-1     EIRASANLAATKM
P.1      EIRASANLAAIKM  T1027I
```

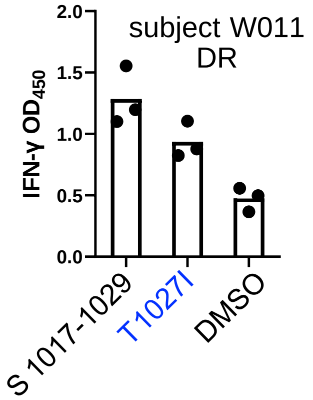

Supplementary Fig. 16

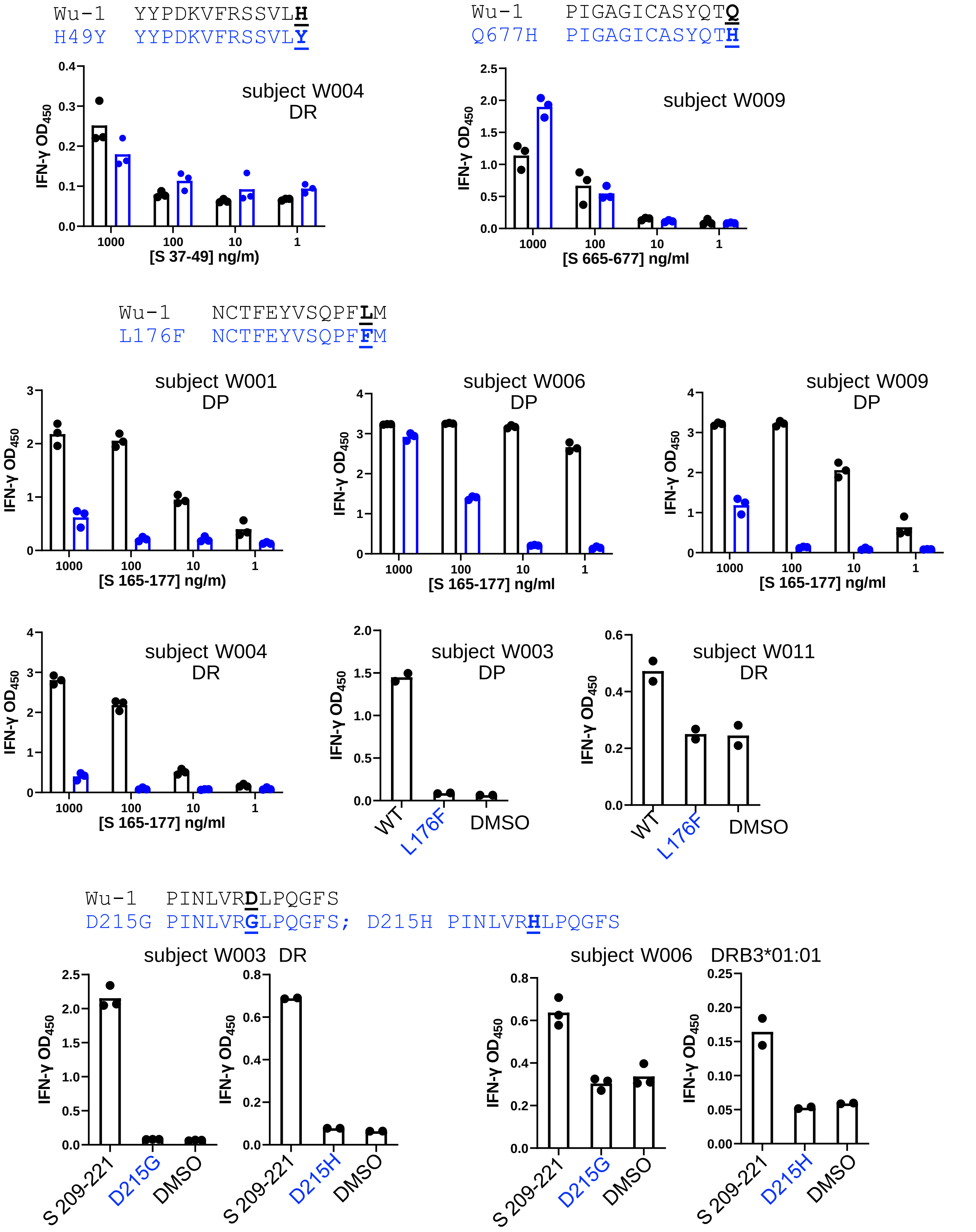

Supplementary Fig. 17

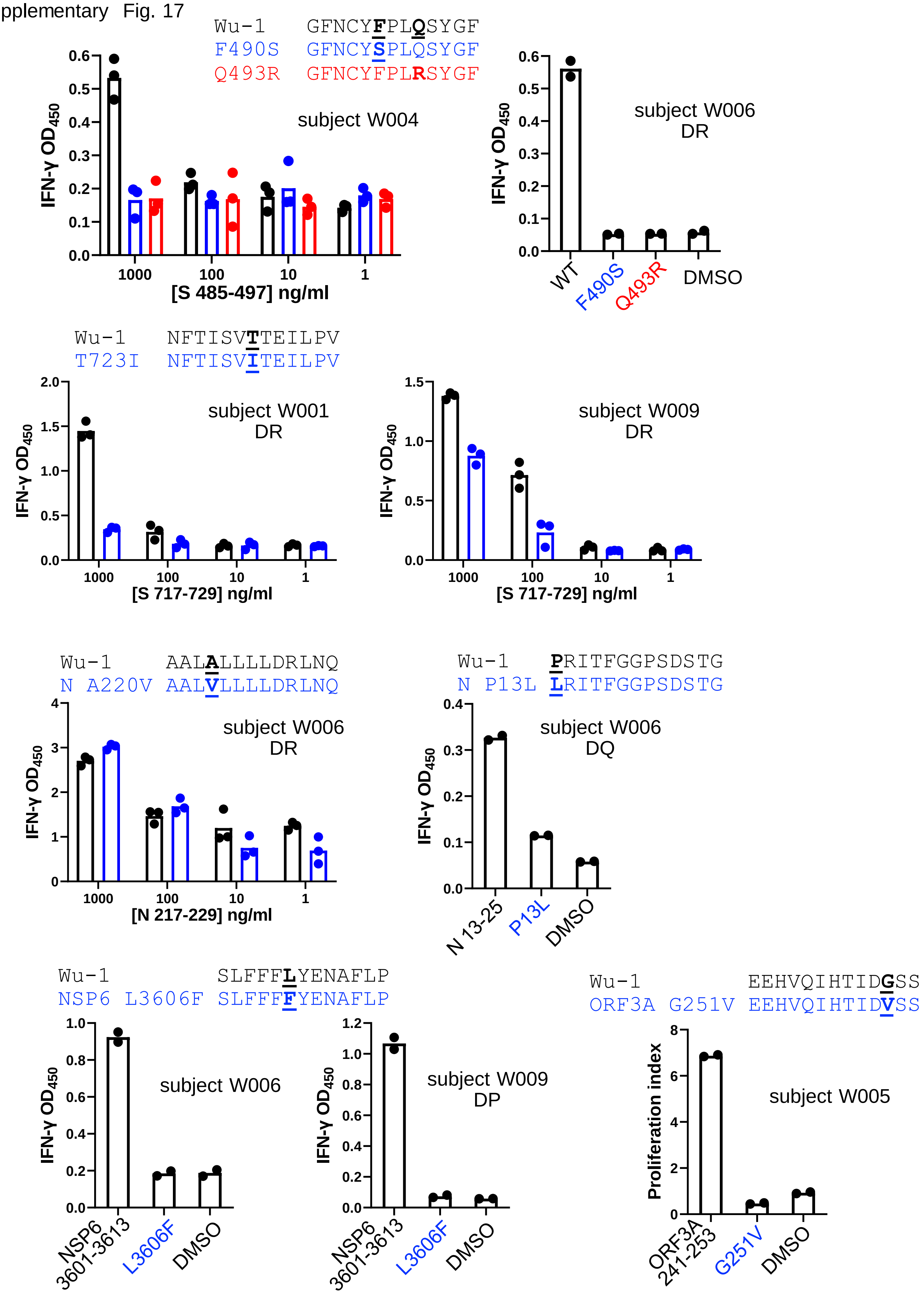

Supplementary Figs. 14-17. CD4 TCL reactivity to variant SARS-CoV-2 peptides. Each graph represents data for one subject and one peptide. The Wu-1 peptide (black) and one or more variant peptides (blue, red) were tested at several concentrations as indicated on the X-axis, or at 1 µg/ml if concentrations are not shown. Variant amino acids are underlined and bolded with shorthand names on the X axis. WT = Wu-1 sequence. DMSO is negative control. Some peptide sets were tested for more than one subject. Subject numbers correspond to Supplementary Table 1. S=spike, N= nucleoprotein. Dots are data from triplicate assays and bars are mean values.
